# Multi-model forecasting of respiratory disease activity in Germany during the 2024–2025 season

**DOI:** 10.64898/2026.07.20.26358471

**Authors:** Johannes Bracher, Daniel Wolffram, Rodrigo Amaral Lind, Nils Bardeck, Micha Böhm, Seba Contreras, Philipp Dönges, Felix Günther, Rolf Kaiser, Jan van de Kassteele, Alexander Kuhlmann, Berit Lange, Barbora Němcová, Viola Priesemann, Ulrich Reinacher, Isti Rodiah, Frank Sandmann, the RESPINOW Study Group, Melanie Schienle

## Abstract

Respiratory diseases cause considerable morbidity in autumn and winter and are a priority in public health monitoring. In Germany, they are subject to a number of surveillance systems, including both pathogen-specific and syndromic indicators. In this paper we present a collaborative multi-target and multi-model real-time forecasting system rolled out during the 2024/25 season, and discuss differences to earlier efforts carried out during the COVID-19 pandemic. We focus on general practitioner consultations for acute respiratory infections (ARI) and hospitalizations for severe acute respiratory infections (SARI), where a total of eight models were run to generate nowcasts and forecasts. In addition, we consider confirmed cases of seasonal influenza and respiratory syncytial virus (RSV) where one forecasting model each was available. As all indicators were subject to retrospective revisions, forecasting models were combined with a nowcasting step. For the SARI and ARI indicators, we combined the available models into an ensemble and conducted detailed quantitative evaluations. Nowcasts overall showed convincing performance, even though for some models Christmas break effects led to an upward bias in early January. Forecasts were overall well-calibrated and most models outperformed simple benchmark models. When taking into account age-stratified targets, most observed performance differences to a persistence baseline were statistically significant. Anticipating the peak timing and magnitude, however, proved to be challenging, with many models predicting too flat curves with a too early turnaround (e.g., in late January rather than mid-February for SARI). The combined ensemble forecast was among the best-performing approaches, but unlike in previous related projects did not consistently outperform individual models. We conclude by discussing learnings on the organization of collaborative forecasting projects in post-COVID-19 times.

**Author summary:** Nowcasting and short-term forecasting are key tasks in epidemic modelling, and have received increasing attention from public health agencies in recent years. So-called Forecast Hubs have become a standard format to structure and coordinate modelling efforts. By collecting time-stamped predictions in standardized formats, they facilitate meaningful model comparisons and the generation of combined ensemble forecasts, which have often been found to be more robust. In this paper, we present the setup of a multi-model and multi-pathogen Forecasting Hub set up in Germany, along with detailed evaluation results for the 2024/25 season. Nowcasting models (i.e., models correcting recent incidence values for reporting delays) showed reliable performance. Forecasting models, on the other hand, mostly outperformed simple reference models, but struggled to anticipate peak timing and magnitude correctly. This paper adds to a growing body of evidence on the real-time performance of epidemic forecasts.

## 1 Introduction

Respiratory diseases are a major source of morbidity and mortality, and are therefore subject to monitoring by public health agencies worldwide. The resulting data streams provide a critical empirical foundation for mathematical modelling of pathogen spread, which for instance during the COVID-19 pandemic played an important role in informing public health decision making (Brooks-Pollock et al., 2021). Models can serve to characterize disease and transmission properties, detect unusual activity, and conceive control strategies, among many other purposes (Keeling and Rohani, 2008). Real-time situational awareness can be supported by model-based nowcasting (the characterization of recent activity based on yet incomplete data; Höhle and an der Heiden 2014) and short-term forecasting (model-based projections for the near future; Held et al. 2017).

In the present paper we report on a collaborative predictive modelling effort conducted in Germany. Starting real-time operations in the 2024/25 season, it extends upon previous efforts (Bracher et al., 2021b; Wolffram et al., 2023) in three major ways. Firstly, we broadened the scope of considered surveillance indicators, covering different syndromes / pathogens and parts of the surveillance pyramid. Specifically, a total of nine models run by five independent research groups were applied to general practitioner consultations for acute respiratory infections (ARI), hospitalizations for severe acute respiratory infections (SARI) and confirmed cases of seasonal influenza and RSV (respiratory syncytial virus; Goerlitz et al. 2021). Secondly, as all indicators were subject to retrospective revisions, nowcasting and forecasting models had to be coupled. A simple approach, examined in a previous retrospective study (Wolffram et al., 2026), consists in computing a bundle of nowcast trajectories, generating forecasts for each of them and pooling these together. This generally enhances the performance of forecasts and thus their utility for monitoring. Lastly, specific attention was given to age-stratified predictions, owing to the fact cases in different age groups may imply different needs for care. Moreover, respiratory disease waves often start and peak earlier in younger age groups (see discussion in Worby et al 2015). A stratified approach may be able to exploit such patterns and thus enhance predictions.

Following good open science practices, real-time forecasts, nowcasts, data and data revisions were systematically collected and visualized in the public *RESPINOW Hub* platform (https://respinowhub.de/; see Supplementary Figure). By default this dashboard showed a combined ensemble forecast computed from all available submissions, which was the main output of the project. The evaluation strategy for the present paper was laid out prospectively in a pre-registration protocol (Bracher and Wolffram, 2024).

The *RESPINOW Hub* is part of a growing ecosystem of *Forecast Hubs* (Reich et al., 2022), which have become a widely used format to coordinate modelling activities. The Forecast Hubs promote good modelling practices such as probabilistic rather than point forecasting, and the parallel operation of several models based on diverse assumptions. The latter notably enables the generation of combined ensemble forecasts, which have often been found to outperform individual models (Reich et al., 2019; Cramer et al., 2022). While early instances of the Hub format stem from the 2010s (e.g., Reich et al. 2019; Johansson et al. 2019), the concept gained traction during the COVID-19 pandemic, most prominently in platforms run by the US (Cramer et al., 2022) and European CDC (Sherratt et al., 2023). In Germany, Hubs have been operated for short-term forecasting of COVID-19 cases and deaths (Bracher et al., 2021b) and nowcasting of hospitalizations (Wolffram et al., 2023) during different phases of the pandemic. After the acute phase of the COVID-19 pandemic, these projects required transitioning to a regime with overlapping seasonal epidemics of various pathogens and changed surveillance practices. Seasonal waves are in principle more predictable than the dynamics of emerging pathogens. This is because pathogen properties are better understood, but also because behavioural feedback loops (Eksin et al., 2019) and non-pharmaceutical interventions (NPIs) do not have a major impact on the course of an outbreak. An important challenge, however, arises from the fact that NPIs perturbed the seasonal dynamics of all respiratory diseases in complex ways over the previous seasons (Ullrich et al., 2021). From a community organization standpoint, modelling collaborations necessarily become smaller in such “times of peace”, and may focus on the consolidation and revision of systems in light of the lessons learned. We return to this aspect in the Discussion section and refer to related works from Italy (Fiandrino et al., 2025), the UK (Kennedy et al., 2026) and Australia (Henderson et al., 2026), as well as the Europe-wide *RespiCast project* (https://respicast.ecdc.europa.eu/; Gozzi et al. 2026).

In line with previous work (Wolffram et al., 2023), we find that nowcasting models were overall reliable, despite some of them temporarily suffering from Christmas break effects. Specifically, difficulties to adjust to changed reporting patterns during the holiday season led to an upward bias in subsequent weeks. Forecasting models were overall well-calibrated, but at the expense of rather wide forecast intervals. The peak magnitude of the ARI and SARI targets was underestimated by all considered models, which expected the turnaround to occur earlier than was observed. Overall most, but not all considered models outperform simple benchmark models. For age-stratified targets, the observed differences are mostly statistically significant, while the smaller number of unstratified population-level incidence targets in most cases does not yield sufficient statistical power. We note that during this first season, not all participating teams were able to provide complete sets of model forecasts over the season in real time. In line with our study protocol we completed certain submissions retrospectively using snapshots of data as available in real time (see Section 4.1 for details). The present study is thus a hybrid of real-time and retrospective model assessment.

The remainder of the article is structured as follows. Section 2 provides an overview of the employed data sources and their particularities. Models and evaluation metrics are introduced in Section 3. In Section 4, we discuss the performance of forecasts qualitatively and quantitatively, before concluding with a discussion in Section 5.

## 2 Data

Respiratory disease activity in Germany is monitored via several different surveillance systems (Goerlitz et al., 2021). While the *RESPINOW* platform is an attempt to integrate as many of them as possible, and displays e.g., virological surveillance data from the National Reference Center (NRZ; Wedde et al. 2025) and the German Clinical Virology Network (Horemheb-Rubio et al., 2022), modelling efforts were focused on four indicators: general practitioner consultations for acute respiratory infections (ARI), hospitalizations from severe acute respiratory infections (SARI), as well as laboratory-confirmed cases of seasonal influenza and RSV. These reflect a desire to cover different levels of the severity pyramid and to balance syndromic and pathogen-specific indicators.

### 2.1 General practitioner consultations for acute respiratory infections (ARI)

The *ARE-Praxis-Sentinel* surveillance system (Goerlitz et al., 2021) consists of more than 500 general practitioners (roughly 1% of all GPs in Germany) who provide information on the number of consultations for respiratory infections. Reporting is done directly to Robert Koch Institute (RKI), typically electronically (SEED-ARE). We use the estimated consultation incidence for acute respiratory infections (ARI; ICD-10 codes J00 – J22, B34.9, and J44.0), which we rescale to absolute count values. Estimates are available in a weekly resolution (with weeks starting on Mondays) and released via RKI’s public GitHub site (https://github.com/robert-koch-institut/) each Thursday. The ARI indicator is not specific to one pathogen and thus forms part of syndromic surveillance. Besides the unstratified national-level time series (00+), we used data on five age groups (0–4, 5–14, 15–34, 35–59, 60+). A graphical display of the time series is available in the top left panel of Figure 1.

**Figure 1:**
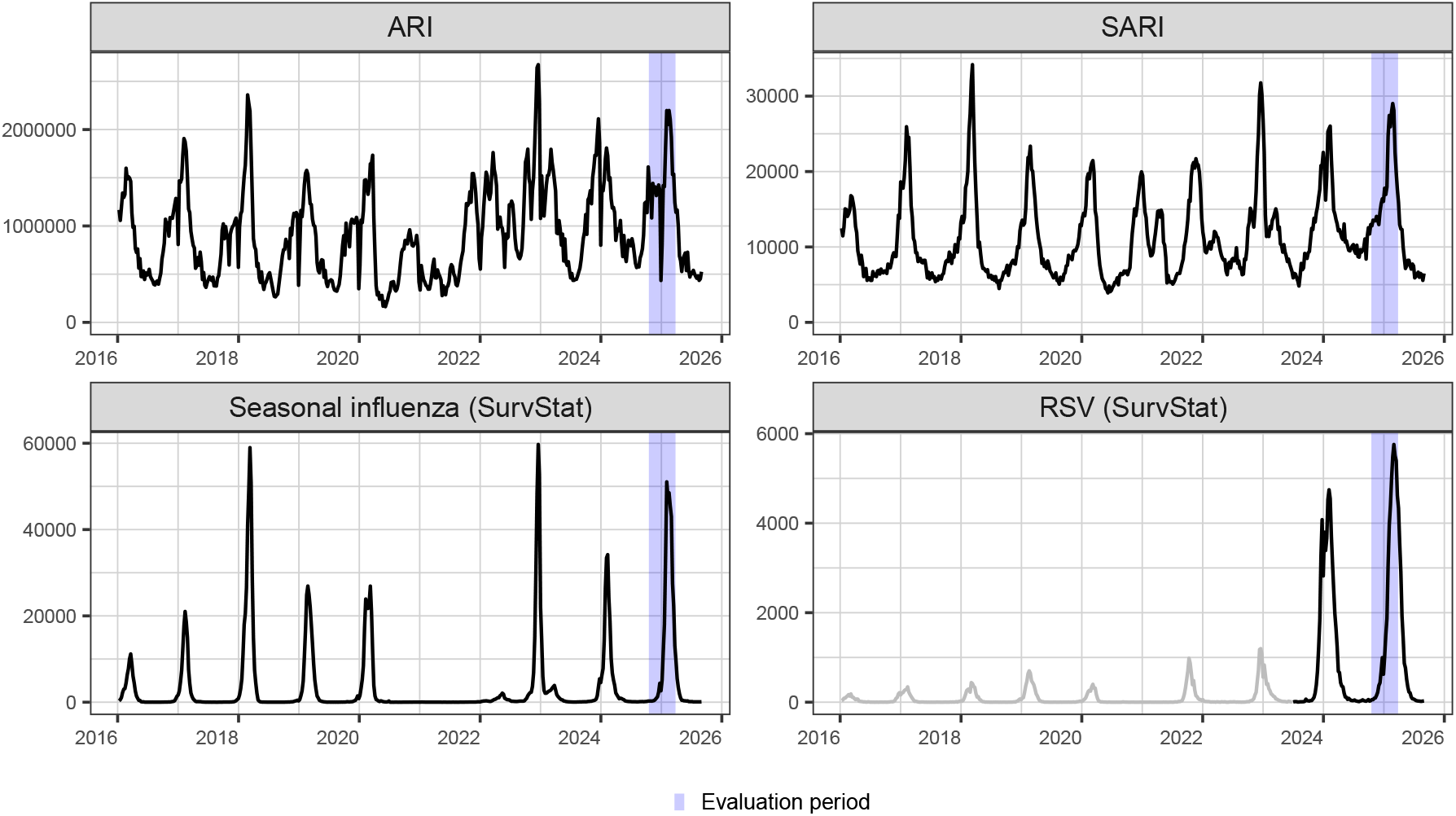
Time series for acute respiratory infections (ARI), severe acute respiratory infections (SARI) and numbers of lab-confirmed cases of seasonal influenza and RSV from the *SurvStat* system. Until 2023, RSV was only notifiable in one German state (Saxony), such that data before this point are greyed out. The purple shaded areas highlight the pre-specified study period discussed in the present paper.

### 2.2 Hospitalizations for severe acute respiratory infections (SARI)

Since fall 2014, RKI has run the *ICOSARI* sentinel system to quantify the incidence of hospitalizations due to severe acute respiratory disease (SARI; Buda et al. 2017; Tolksdorf et al. 2022). The sentinel consists of roughly 70 hospitals (accounting for 5–6% of all hospitalizations in Germany) and the SARI case definition is based on a set of ICD-10 diagnostic codes (J09–J22); see Buda et al. (2017) for details. The age stratification is slightly more detailed, with an extra split into groups 60–79 and 80+. As for ARI consultations, we re-scale the numbers published as incidences per 100,000 population to absolute counts.

### 2.3 SurvStat

For a large number of communicable diseases, laboratory-confirmed cases are notifiable under the *German Infection Protection Act*. Local health authorities (*Gesundheitsämter*) receive reports from general practitioners and laboratories, and via state-level administrations (*Landesbehörden*) forward them to RKI. The resulting data are made available in aggregate format via the *SurvStat* system (https://survstat.rki.de/). We here consider data on RSV and seasonal influenza, which are available by state and age group. We note that until the 2022/23 season, RSV was only notifiable in the state of Saxony, explaining why the reported absolute numbers shift by an order of magnitude in 2023. As the data from Saxony informed model fits, we display them in Figure 1, but grey them out to avoid confusion.

It should be noted that laboratory analyses are only performed for a fraction of all patients seeking care, meaning that *SurvStat* only covers part of the actual disease incidence. Reporting completeness can vary over time and depends on many aspects, including healthcare seeking behaviour and reimbursement policies. While trends are thus interpretable within each season, comparisons across seasons are not straightforward.

### 2.4 Data revisions

A characteristic of the ARI and SARI indicators is that recent values are subject to revisions, which needs to be taken into account in our forecasting tasks. The patterns of revisions, however, differ. SARI hospitalizations are almost exclusively corrected upwards, as revisions result from the *delayed addition* of events to the record after the first data release. We illustrate this for the SARI data in the left panel of Figure 2. Typically, roughly 75% of all hospitalizations for a given week are contained in the initial data release, the remainder is added over subsequent weeks (most of them with a delay of just one week).

**Figure 2:**
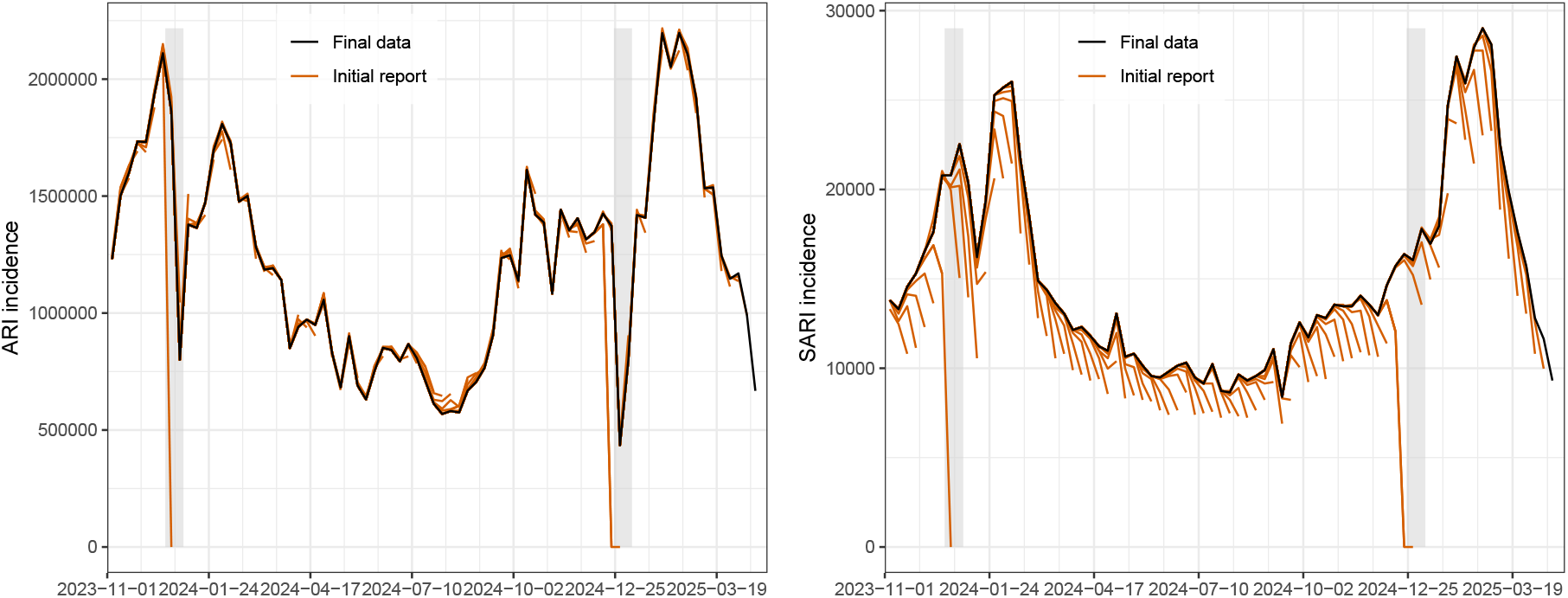
Revisions of the ARI (left) and SARI (right) time series. Black lines show the final consolidated version of the time series, while the overlaid orange lines show the versions published in different weeks. Grey shaded areas show the Christmas / New Year period (last and first calendar week of the year).

For the ARI indicator, revisions are overall less pronounced, as can be seen from the middle panel of Figure 2. Revisions are more symmetric and commonly occur both upward and downward. This is explained by a different revision mechanism, namely *revised averages*. The ARI numbers result from a weighted average of numbers from different sentinel sites, which are re-computed whenever information from certain sentinel sites is added with a delay. Revisions are therefore largely unsystematic and manifest as fluctuations rather than consistent upward corrections.

In the *SurvStat* data, revisions are typically upwards, but are negligible in many weeks (shown for influenza in Supplementary Figure S6).

## 3 Methods

### 3.1 Definition of forecasting tasks

In each week of our study period (submission dates 17 October 2024 – 24 March 2025, submissions made during the weeks of Christmas and New Year excluded), we collected nowcasts and forecasts, which are based on data releases as available on Thursday (the day when SARI and ARI data are updated). Nowcasts refer to *horizons h* = *−*3, *…*, 0 weeks, with *h* = 0 denoting the week ending on the Sunday preceding the data release. Forecasts are issued for horizons *h* = 1, *…*, 4. The prediction target consists in the weekly numbers of consultations, cases or hospitalizations reported with a delay of at most 4 weeks, which in practice is very close to the respective total numbers (see Wolffram et al. 2023 for a more detailed discussion). Forecasts are collected for weekly totals as well as per age group, and stored in a standardized quantile format (with quantile levels 2.5%, 10%, 25%, 50%, 75%, 90%, and 97.5%). This follows conventions from previous Forecast Hub applications (e.g., Cramer et al. 2022, Wolffram et al. 2023).

### 3.2 Evaluation metrics

We evaluate forecasts using the *weighted interval score* (WIS, Bracher et al. 2021a), which is a quantile-based approximation of the widely used continuous ranked probability score (CRPS; Gneiting and Raftery 2007). Given quantiles *q*_1_, *q*_2_, *… q*_*K*_ at levels *τ*_1_ *< τ*_2_ *< … < τ*_*K*_ *∈* (0, 1) and an observed value *y* it is computed as

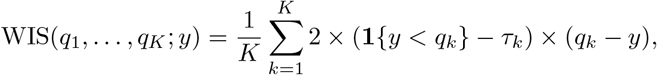

with **1** the indicator function. The WIS is a probabilistic generalization of the absolute error and is thus negatively oriented (lower values represent better performance). To put WIS averages into perspective, we also report relative WIS values (Cramer et al., 2022), given by average WIS of a given model

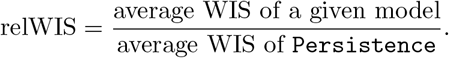

The use of `Persistence` (rather than `Historical`) as the baseline was not pre-registered, but was made based on conventions in the recent literature (Cramer et al., 2022; Sherratt et al., 2023; Gozzi et al., 2026). To assess the statistical significance of observed differences against this baseline, we employ permutation tests as implemented in the R package `surveillance` (Meyer et al., 2017). Scores for the same submission date and stratum but for different horizons are permuted jointly to reduce correlations between scores.

WIS values and their averages can be decomposed into components for forecast spread, overprediction, and underprediction (Bracher et al., 2021b), which we will use to enhance the interpretability of performance summary plots. Put shortly, for an individual WIS value, the spread component corresponds to the smallest WIS value which can be achieved with a given set of predictive quantiles (for symmetric quantile levels, this is the WIS value achieved if the observation equals the predictive median). The difference between the achieved WIS and this minimum possible value is counted as the overprediction or underprediction component, depending on whether the observation fell above or below the predictive median. See also Resin et al. (2026, Section 2.3) for the theoretical underpinnings.

While the WIS applied to incidence counts is our pre-registered main evaluation metric, its scale dependence implies that average scores tend emphasize weeks and strata with high incidences (Bracher et al., 2021a). This notably means that getting the peak right is much more important than e.g., correctly anticipating the season onset. An alternative which gives roughly equal weights across weeks and strata is the application of the WIS to a transformed target of log(incidence + 1) (Bosse et al., 2023). We will report this version as a secondary metric.

In addition, we evaluate point forecasts using absolute errors, and forecast calibration using coverage proportions of 50% and 95% prediction intervals. The coverage proportion is the fraction of cases in which a prediction interval successfully covered the observation. We note that similarly to Wolffram et al. (2023), and in line with our study protocol, we avoid the problem of missing submissions by using retrospective fill-in predictions. See Section 4.1 for details.

### 3.3 Nowcasting and forecasting methods

We consider a total of nine models, with Table 1 summarizing which of them cover which indicators. The labels here consist of the research team running the model and the model name, but we will mostly omit the team name for brevity in the remainder of the paper. Three broad model categories can be distinguished.

**Table 1:** Models contributed to the *RESPINOW* Hub and targets they cover. The column age stratification indicates whether forecasts per age group were generated.

| **Nowcasting** |  |  |  |  |  |
|---|---|---|---|---|---|
| **Model** | SARI | ARI | Influenza | RSV | age stratification |
| KIT-simple_nowcast | ✓ | ✓ | ✓ | ✓ | ✓ |
| KIT-epinowcast | ✓ |  | ✓ |  | ✓ |
| RIVM-GAM | ✓ |  | ✓ |  | ✓ |
| RKI-Pilot_01 | ✓ |  |  |  | ✓ |
| **Forecasting** |  |  |  |  |  |
| Model | SARI | ARI | Influenza | RSV | age stratification |
| KIT-hhh4 | ✓ | ✓ |  |  | ✓ |
| KIT-LightGBM | ✓ | ✓ |  |  | ✓ |
| KIT-TSMixer | ✓ | ✓ |  |  | ✓ |
| MPIDS-PS_embedding | ✓ | ✓ |  |  |  |
| HZI-ODEmodel |  |  | ✓ | ✓ |  |

#### Nowcasting models

`KIT-simple_nowcast` (Wolffram et al., 2026; Johnson et al., 2026), `KIT-epinowcast` (Abbott et al., 2026), `RKI-Pilot_01` (Günther et al., 2025) and `RIVM-GAM` van de Kassteele et al. (2019) are pure nowcasting models and thus only applied to horizons *h* = *−*3 through 0 weeks. We note that only `KIT-simple_nowcast` is applicable to the *revised average* setting encountered for ARI, while the two other methods can only be applied to the SARI and SurvStat indicators.

- **Mechanistic models for pathogen-specific indicators:** `HZI-ODEmodel` is a classic compartmental (SIR-type) model and applicable to the *SurvStat* influenza and RSV targets.
- **Data-driven statistical models for syndromic indicators:** Mechanistic models are challenging to adapt to multi-pathogen syndromic indicators. However, as these time series are characterized by strong autocorrelations and rather stable seasonal patterns, four data-driven statistical models were applied to the ARI and SARI data.
  – `KIT-hhh4` is a seasonal count-time series model from the endemic-epidemic class (Bracher and Held, 2022).
  – `KIT-LightGBM` is an application of the widely used LightGBM gradient boosting approach (Ke et al., 2017).
  – `KIT-TSMixer` is an application of the TSMixer library (Chen et al., 2023), a neural network tailored to time series forecasting. Both `KIT-LightGBM` and `KIT-TSMixer` contain lagged incidence counts and the calendar week as features. They moreover generate predictions for all age groups simultaneously and can hence exploit relationships betwee the different time series; see Wolffram et al. (2026) for details on these three models and their implementation.
  – `MPIDS-PS_embedding` matches the observed incidence curve to historical ones in a multidimensional phase space and generates forecasts based on a set of closest matching curves. Details are provided in Supplementary Section B.2.

All `KIT` models use the `KIT-simple_nowcast` as an input to account for reporting delays, while `MPIDS-PS_embedding` has an integrated correction step.

In addition, two benchmark models were used to put performance into context, and two different ensembles were computed (see Wolffram et al. 2026 on implementation details). Lastly, for ARI, an ensemble forecast from the European *RespiCast* platform was available, which we include in our comparison.

- `Persistence` is a flat-line forecaster, with uncertainty intervals obtained from past prediction errors. It includes a nowcasting step (based on the `simple_nowcast`) in order to avoid a downward bias for SARI.
- `Historical` is a purely seasonal benchmark, where predictive distributions are obtained from past observations in the same and neighbouring calendar weeks of previous years.
- `KIT-ensemble_complete` is a quantile-wise average (mean) of all member models (excluding benchmark models). `KIT-ensemble_realtime` is an ensemble limited to those models that were available in real time (see Section 4.1 on which models were available in which week). This version is only available for the SARI target, which was the only one with sufficiently many member models computed in real time.
- `RespiCast-Ensemble` is the ensemble forecast from the European project *RespiCast* (Gozzi et al. 2026; available from the repository https://github.com/european-modelling-hubs/RespiCast-SyndromicIndicators). It is only available for the total ARI incidence (without age stratification) and is computed from 7–10 member models, depending on the exact week.

The quantile averaging approach used for our ensembles, commonly referred to as *Vincentization* (Genest, 1992), is more practical than e.g., the linear pool when forecasts are collected as quantiles. It is therefore widely used in the Forecast Hub ecosystem; see Ray et al. (2023) for a comparison of different approaches and extensions to weighted ensembles.

## 4 Results

### 4.1 Model availability in real time

During the first operational season of the RESPINOW Hub, for a variety of reasons not all teams were able to consistently provide real-time submissions. To nonetheless provide a comprehensive evaluation of the realtime potential of different models we therefore completed the sets of forecasts with retrospective submissions wherever necessary (Bracher and Wolffram 2024; see also Henderson et al. 2026 for a similar approach). These were consistently based on data versions as available in the respective week of forecast submission, facilitated by the history of our GitHub repository and guidance by the organizers. As information leakage nonetheless may occur, we summarize which submissions were added retrospectively in Figure 3.

**Figure 3:**
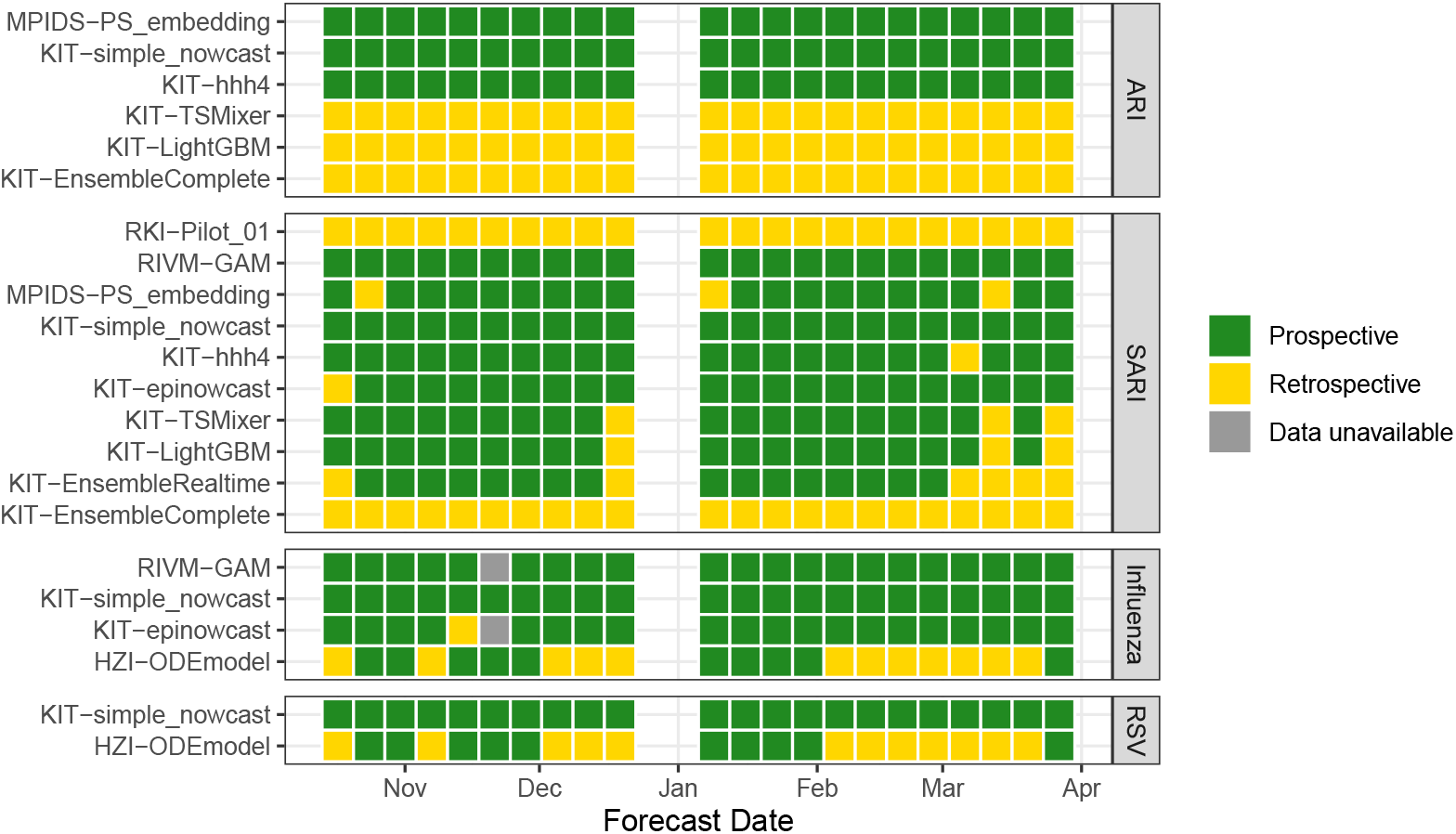
Completeness of submissions to the *RESPINOW* Hub by target and week of the study period.

### 4.2 Qualitative assessment of forecasts

To provide an intuition for the behaviour of our nowcasts and forecasts, we present several types of visual displays. These enable pinpointing issues encountered for different models which will help making sense of our quantitative performance assessment in Section 4.3.

#### 4.2.1 Visual inspection of ensemble forecasts

We first show a set of point predictions and uncertainty intervals as they would be provided to a user of the *RESPINOW* Hub. These are shown with consolidated time series as well as the respective versions available at the time predictions were issued.

##### ARI

The left column of Figure 4 shows `ensemble_complete` forecasts for ARI, with three panels to show a larger set of forecast dates. Data revisions were overall minor during this period (little discrepancies between initial data shown in red and consolidated data shown in black). It is therefore of little surprise that the nowcasts exhibited narrow uncertainty intervals (blue, barely distinguishable) and closely matched the target time series. Point forecasts (green lines) were well-aligned with the observed data during the early and late phases of the season, but the drop in the Christmas weeks was not well reflected despite its recurrent nature (see Figure 1). The following steep increase was within the uncertainty bounds, but likewise not well-anticipated. Overall, the uncertainty intervals were rather wide, with even 50% prediction intervals covering the observations in most cases.

**Figure 4:**
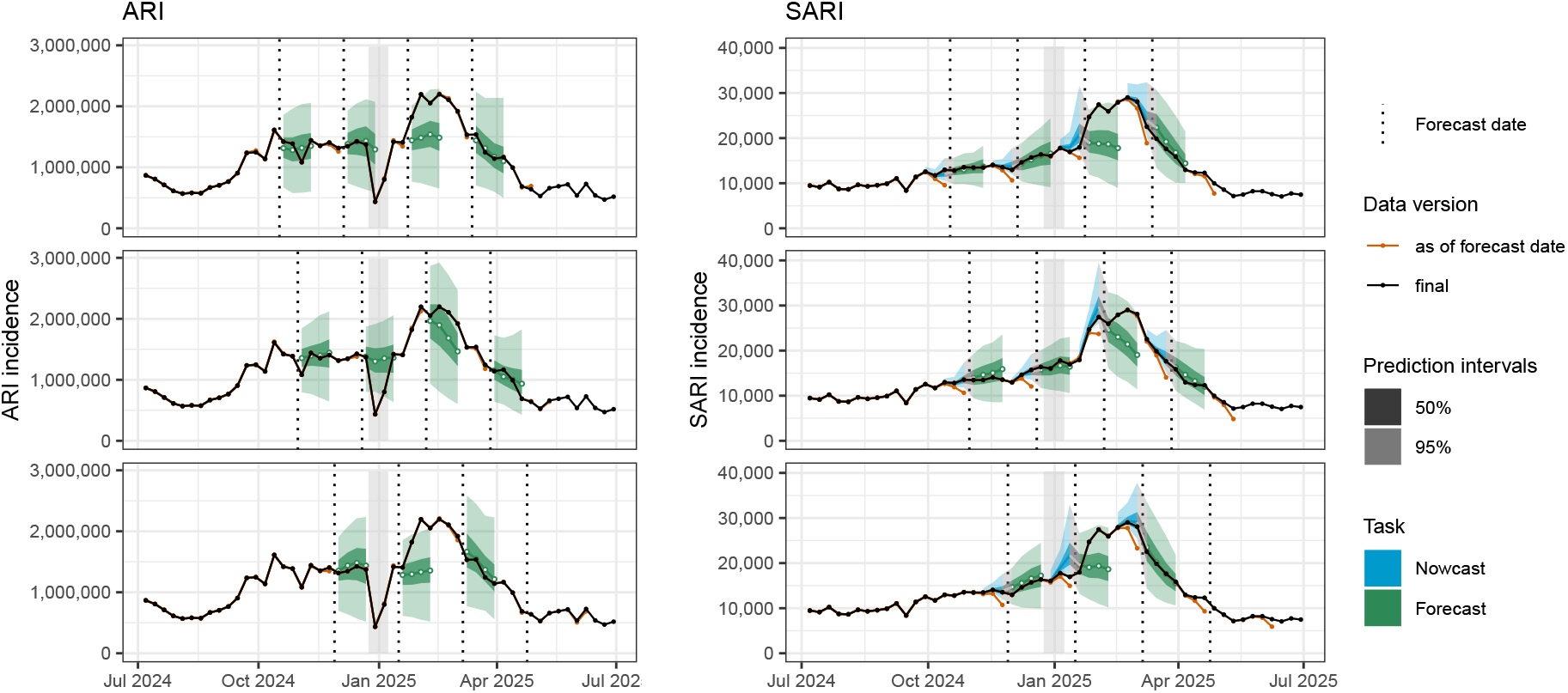
Selected `simple_nowcast` nowcasts and `EnsembleComplete` forecasts for the ARI (left column) and SARI (right) indicators. Three panels are used for each indicator in order to show a larger number of trajectories. Vertical lines show times at which predictions were issued, with nowcasts (blue) to the left of the line and forecasts (green) to the right. Data as available in the respective week are shown in orange. Nowcasts and forecasts are displayed as predictive medians (lines) along with shaded areas for 50% (dark) and 95% (light) uncertainty intervals. Grey shaded areas show the Christmas / New Year period. A second plot showing the remaining weeks can be found in Supplementary Figure S7.

##### SARI

The SARI time series being subject to more pronounced revisions, this indicator is of main interest for the nowcasts (shown in blue). Overall, these agreed well with the consolidated values, with the exception of some over-prediction in January (bottom right panel in Figure 4). A more detailed discussion is provided in Section 4.2.2. SARI has clearer seasonal patterns without pronounced Christmas effects, leading to somewhat better-behaved forecasts with narrower uncertainty intervals. The peak timing, which is known to be hard to predict, was anticipated too early, while the early and late phase of the season were again well-predicted.

##### SurvStat seasonal influenza and RSV

Forecasts for the influenza and RSV targets were only available in real time for 10 out of 22 weeks (Figure 3). The mechanistic `ODEmodel` had to be executed manually each week, and unlike the fully automated statistical pipelines for the syndromic indicators required some continued manual tuning and diagnostic checking. Retrospectively completing missing weeks would have required an analyst to run the model with full knowledge of the season’s subsequent course, introducing a risk of hindsight bias. We therefore focus on a descriptive assessment of the weeks covered in real time, which covered some key moments of the season. These are shown in Figure 5. For influenza, the typical steep increase occurred earlier than anticipated by the model. While uncertainty intervals were overall too narrow, forecasts produced 3-4 weeks before the peak were relatively accurate, predicting epidemic turnover with roughly correct timing. For RSV, on the other hand, the season onset was predicted well, but peak timing was predicted too early. Uncertainty intervals were considerably wider, with the lower ends of 95% intervals often reaching almost zero. As a consequence, coverage of the observations by the prediction intervals was almost complete.

**Figure 5:**
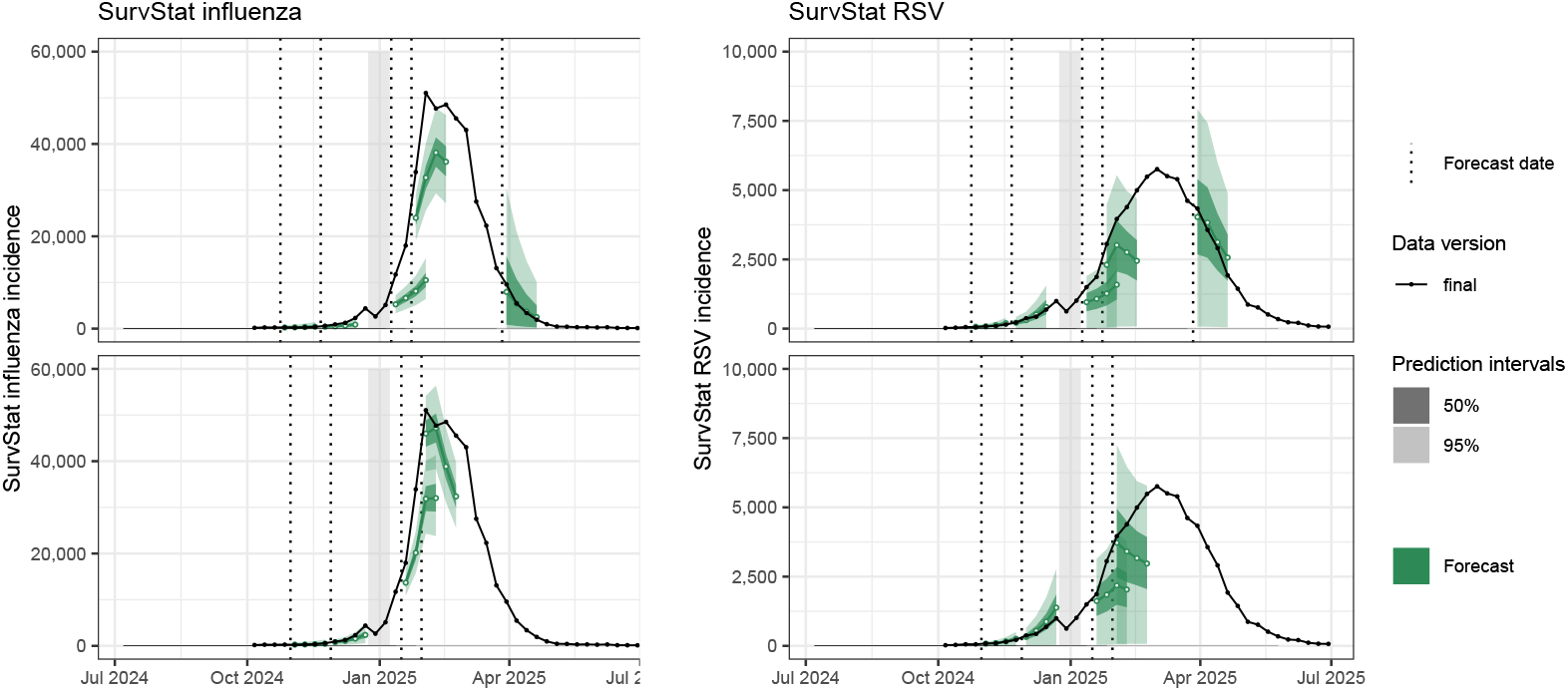
Selected nowcast and forecast trajectories for the SurvStat influanza (left column) and RSV (right) indicators. Plot elements are as described for Figure 4. Note that data revisions are negligible for these indicators (see Supplementary Figure S6) and hence not displayed here.

#### 4.2.2 Same-week nowcasts from different models

We next provide a more comprehensive perspective on the behaviour of our different nowcasting models. We here focus on the SARI target, where nowcasting is more relevant, and the hardest task of same-week nowcasts (horizon *h* = 0 weeks). Figure 6 displays these for each week of our study period, overlaid with the initially reported values available at the time of nowcast. The `RIVM-GAM` and `RKI-Pilot` models were overall well-behaved, with almost complete coverage of 95% prediction intervals. The `simple_nowcast` and `EpiNowcast` models worked well up to the Christmas break, but subsequently suffered from an upward bias and (especially for `EpiNowcast`) increased nowcast variability. This is explained by the fact that neither had a mechanism to account for the different (much longer) delays for calendar weeks 52 and 1. This led to an upward bias in nowcasts for the following weeks. For `simple_nowcast`, a fix was made in week 3, leading to a return to good predictive performance. For `EpiNowcast`, point nowcasts returned to a reasonable behaviour without intervention, but uncertainty intervals remained excessively wide.

**Figure 6:**
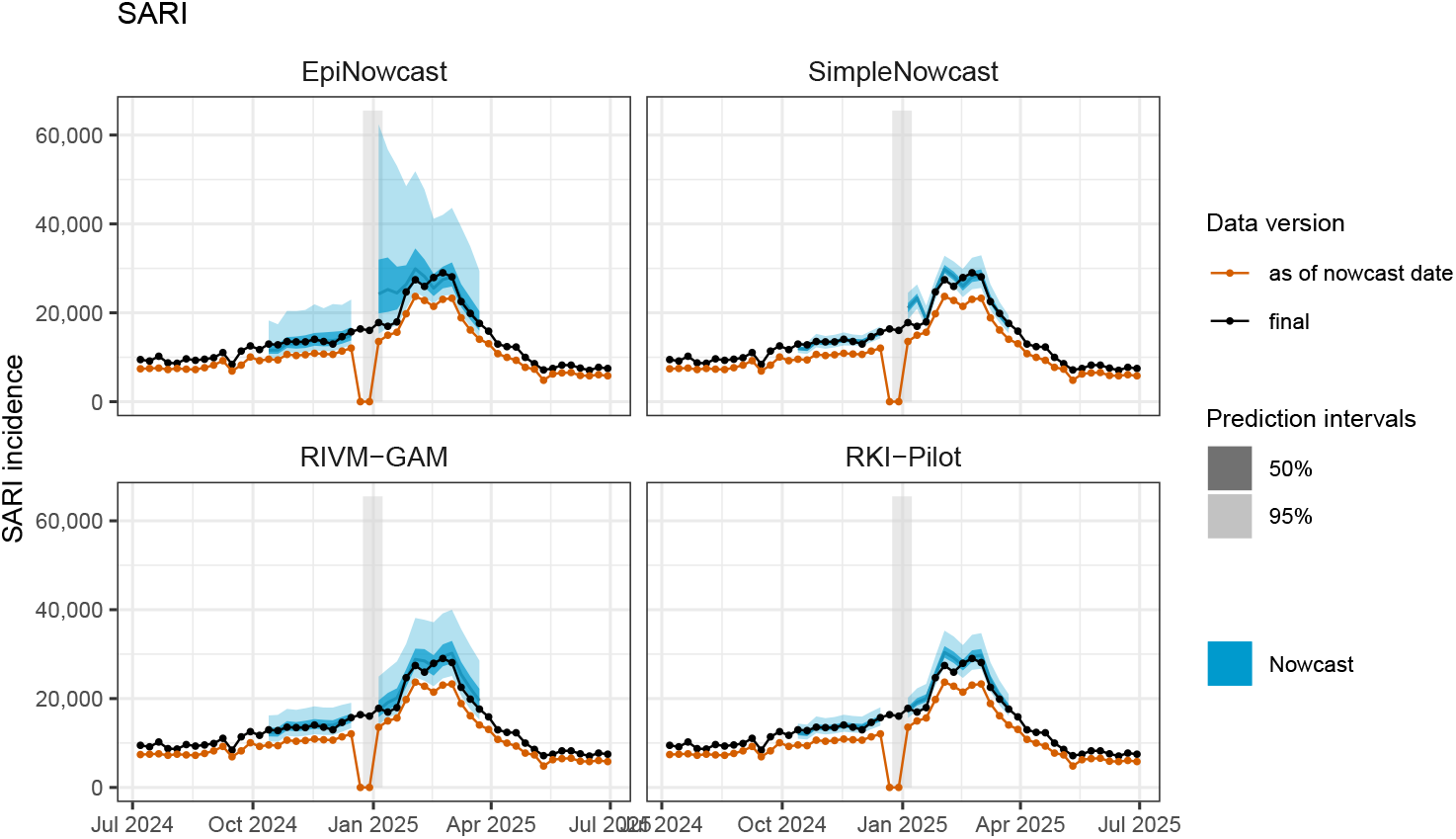
Same-week nowcasts for SARI from the four different models. Grey shaded areas show the Christmas / New Year period

#### 4.2.3 Point forecasts from different models

Detailed displays of probabilistic nowcasts and forecasts as in Figure 4 quickly run into overplotting issues and are limited in how many forecasts can be compared simultaneously. We therefore adopt a plot type from Paireau et al. (2022), where point forecast trajectories are shown per week and model. Figure 7 shows the result for the ARI indicator. The two issues identified for the ensemble forecasts in Section 4.2.1 – overprediction during the Christmas period and premature prediction of the peak – turn out to be shared by most member models. `LightGBM` and the `RespiCast-Ensemble` (which was not produced by the *RESPINOW* collaboration) incorrectly predicted an increase early in the season. `LightGBM` then continued to produce the most reasonable-looking forecasts from December onward, with signs of the Christmas dip (likely thanks to the inclusion of the calendar week as a feature). It predicted peak timing, though not height, roughly correct. `hhh4` throughout the season only predicted very moderate increases or decreases, which we retrospectively found to be a consequence of not explicitly accounting for the Christmas dip (see Appendix B.1 for details). Indeed, a simple extension allowing the model’s autoregressive parameters to be different in the first and last calendar week of the year leads to a remarkable improvement of the model (see panel added in the bottom row of Figure 7). This suggests that the other models, too, could plausibly be improved by providing the Christmas break as an explicit feature.

**Figure 7:**
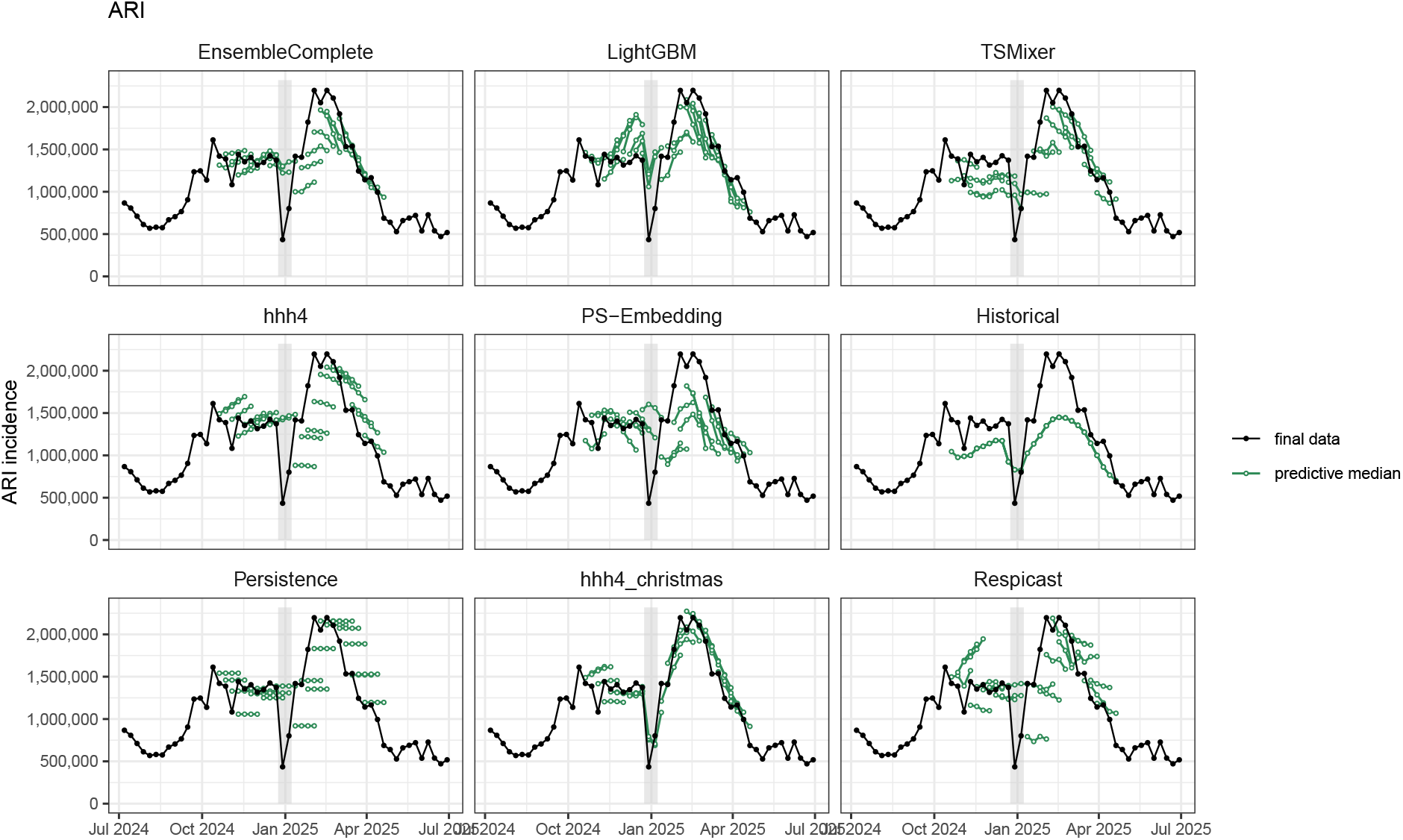
Predictive medians from the ensemble, individual and baseline models for the ARI indicator. An additional retrospective variation of the `hhh4` model has been included (`hhh4_christmas`); see main text and Appendix B.1 for details. Grey shaded areas show the Christmas / New Year period

The corresponding plot for the SARI target can be found in Figure 8. Here, `LightGBM`, `TSMixer` and `PS_Embedding` show downward biases for much of the season and premature predictions of the peak. The `hhh4` model fares somewhat better, but still predicts peak too early. The `RespiCast-Ensemble` is not available for this time series.

**Figure 8:**
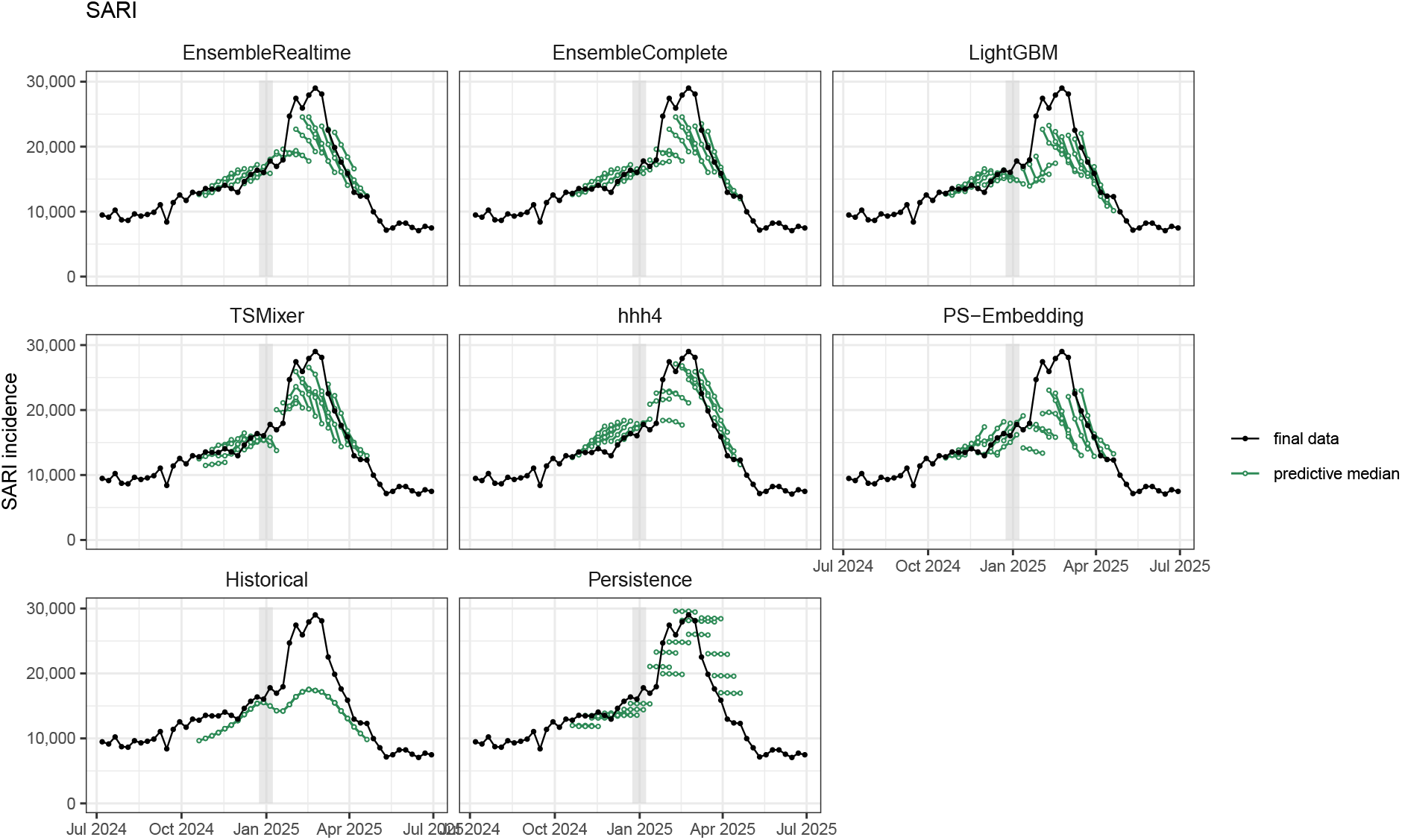
Predictive medians from the ensemble, individual and baseline models for the SARI indicator.

### 4.3 Formal forecast evaluation

Following these qualitative observations, we move to a more formal assessment focused on the ARI and SARI targets.

#### Interval coverage

We first assess calibration in terms of prediction interval coverage. Figure 9 summarizes this for the ARI and SARI targets. For the ARI target coverage is overall good, though 95% prediction interval coverage is mostly below nominal (most pronounced for `PS_Embedding`). For SARI, the forecast models are likewise well-calibrated, while among the nowcast models only the `RKI-Pilot` model achieves nominal coverage. This pattern is even more pronounced for the age-group wise targets (Supplementary Figure S11), and the remaining nowcasting models show substantial under-coverage. As we show in Supplementary Figures S12 and S13, however, the coverage issues arise mainly for horizons *−*3 and *−*2 weeks, which are of limited practical interest as the data are almost complete at this point. For the most relevant same-week nowcasts, coverage is close to nominal for 95% intervals, with `EpiNowcast` and `RIVM-GAM` even over-covering. 50% intervals are conservative for all considered models. This agrees with the visual impression from Figure 6.

**Figure 9:**
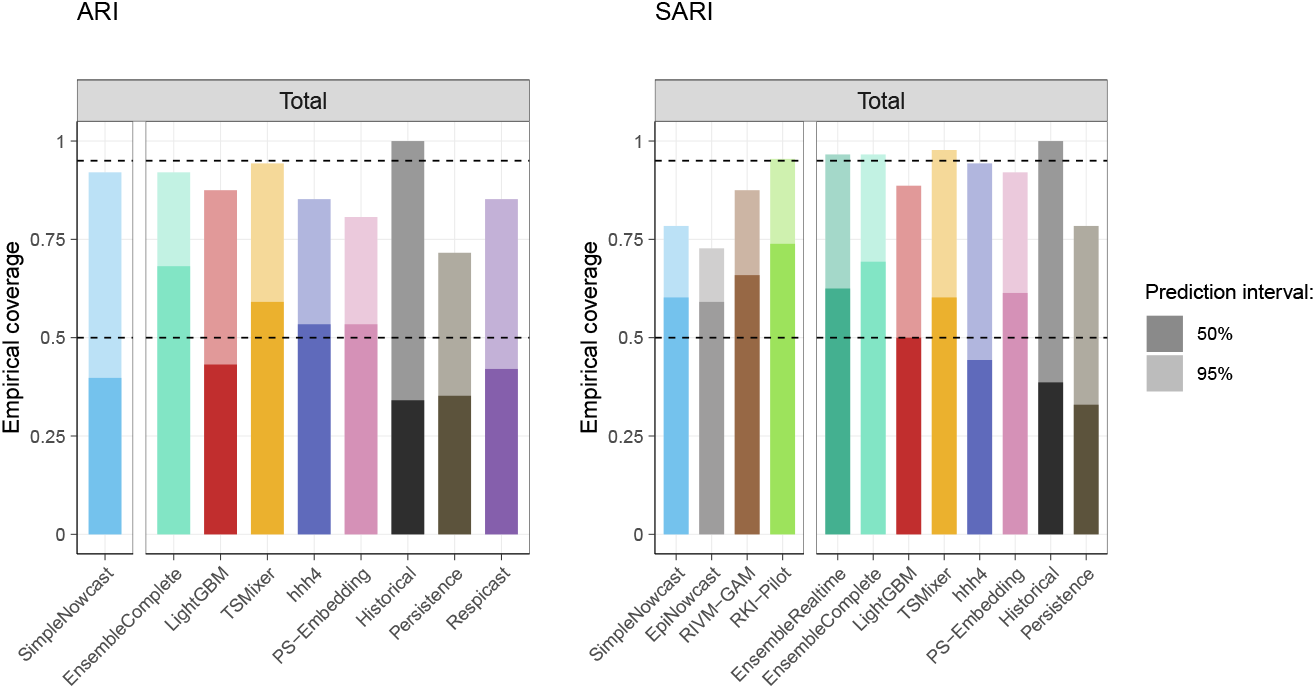
Coverage of 50% (dark shaded) and 95% prediction intervals (light) for the ARI (left) and SARI (right) targets. Fractions are averaged across forecast horizons and weeks. Within each panel, results for nowcasting and forecasting models are shown separately. Results for age-group specific nowcasts and forecasts are shown in Supplementary Figure S11 and qualitatively similar.

#### Score-based evaluation

We summarize mean WIS and absolute errors for ARI and SARI in Figures 10 and 11, respectively. Numerical values can be retrieved from Supplementary Tables S1 and S2, respectively. The top left panels in each figure refer to nowcasts and forecasts for the total incidence (pooled across age group), while the top right refers to the performance for age-group specific time series. As WIS is a scale-dependent metric, it is expected that average scores for the age-group wise forecasts are lower. For ARI, the ensemble model and two member models (`LightGBM` and `TSMixer`) achieve moderate improvements over the two benchmark models. The other members (`hhh4` and `PS_Embedding`) as well as the `RespiCast-Ensemble` are on par with the `Persistence` baseline and slightly outperformed by `Historical`. A similar picture can be found for SARI, though `hhh4` fares better than `LightGBM` in this case. Among the nowcasting models, the (retrospectively applied) `RKI-Pilot` performs best; here it should be noted that scores for nowcasting and forecasting models cannot be compared as they refer to different horizons (−3 through 0 for nowcasting, 1 through 4 for forecasting). Similarly to findings by Moss et al. (2026), but unlike in many other previous efforts, the ensemble forecasts do not clearly outperform the member models. For SARI, the difference between the ensemble forecasts from all models (`Ensemble-Complete`) and only models available in real time (`EnsembleRealtime`) is very similar.

**Figure 10:**
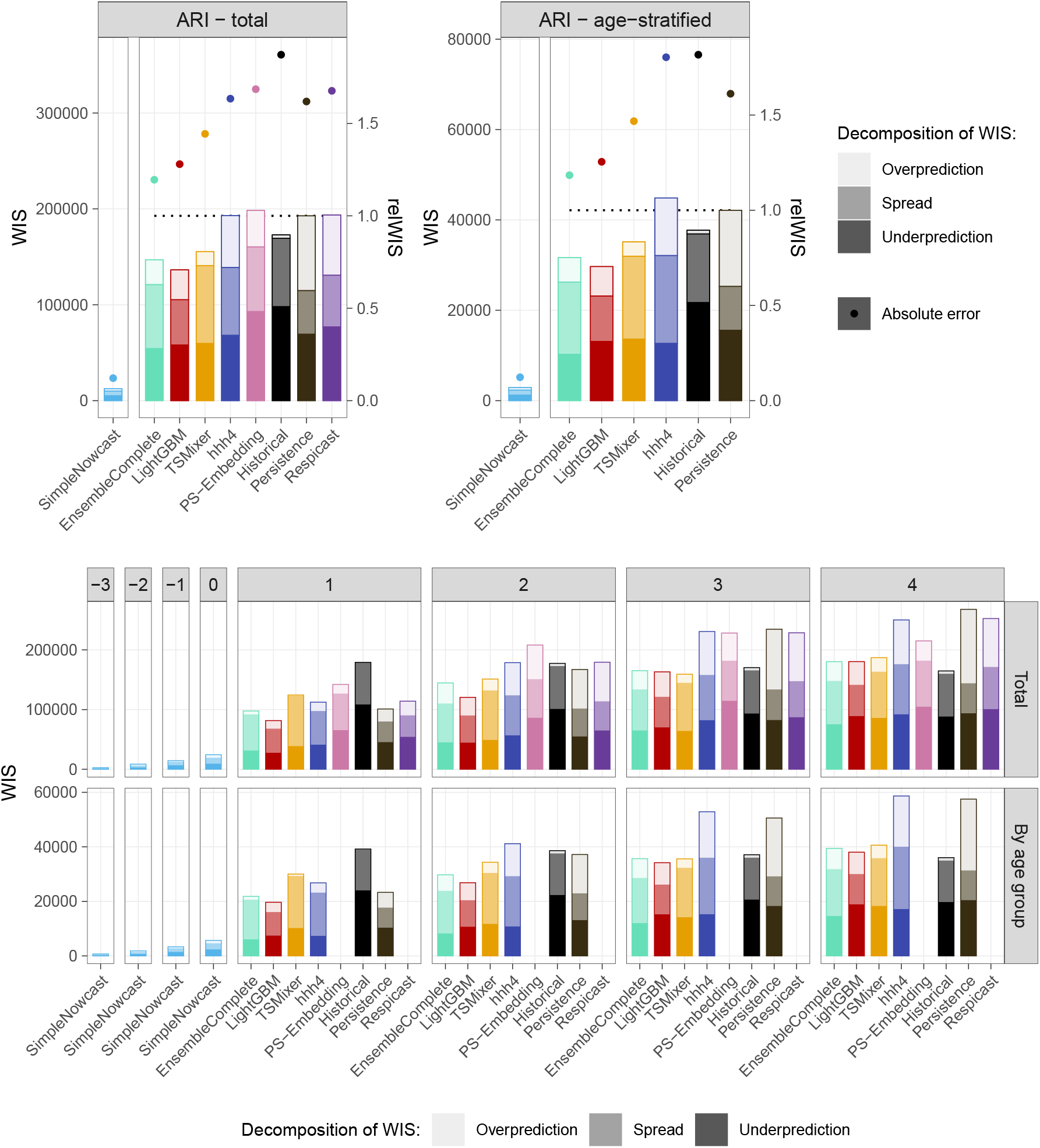
Summary of forecast performance for ARI in terms of weighted interval scores and absolute errors. Top left: Score averages for the predictions of weekly totals (across horizons and forecast dates). Top right: Score averages for age-group specific predictions. Note that results from nowcast and forecast models are visually separated as they refer to different sets of horizons. In the forecast panel, the dotted horizontal line indicates the mean WIS of the `Persistence` baseline, and the second axis to the right shows relative WIS with respect to this baseline (i.e., the ratio of the mean WIS achieved by a model and the average WIS of the baseline). Bottom: Stratified results by nowcast / forecast horizon. We here omit absolute errors to reduce y-axis limits. Average WIS values stratified by age group are available in Supplementary Figure S8. A numerical summary of the displayed values, along with statistical significance tests, is given in Supplementary Table S1.

**Figure 11:**
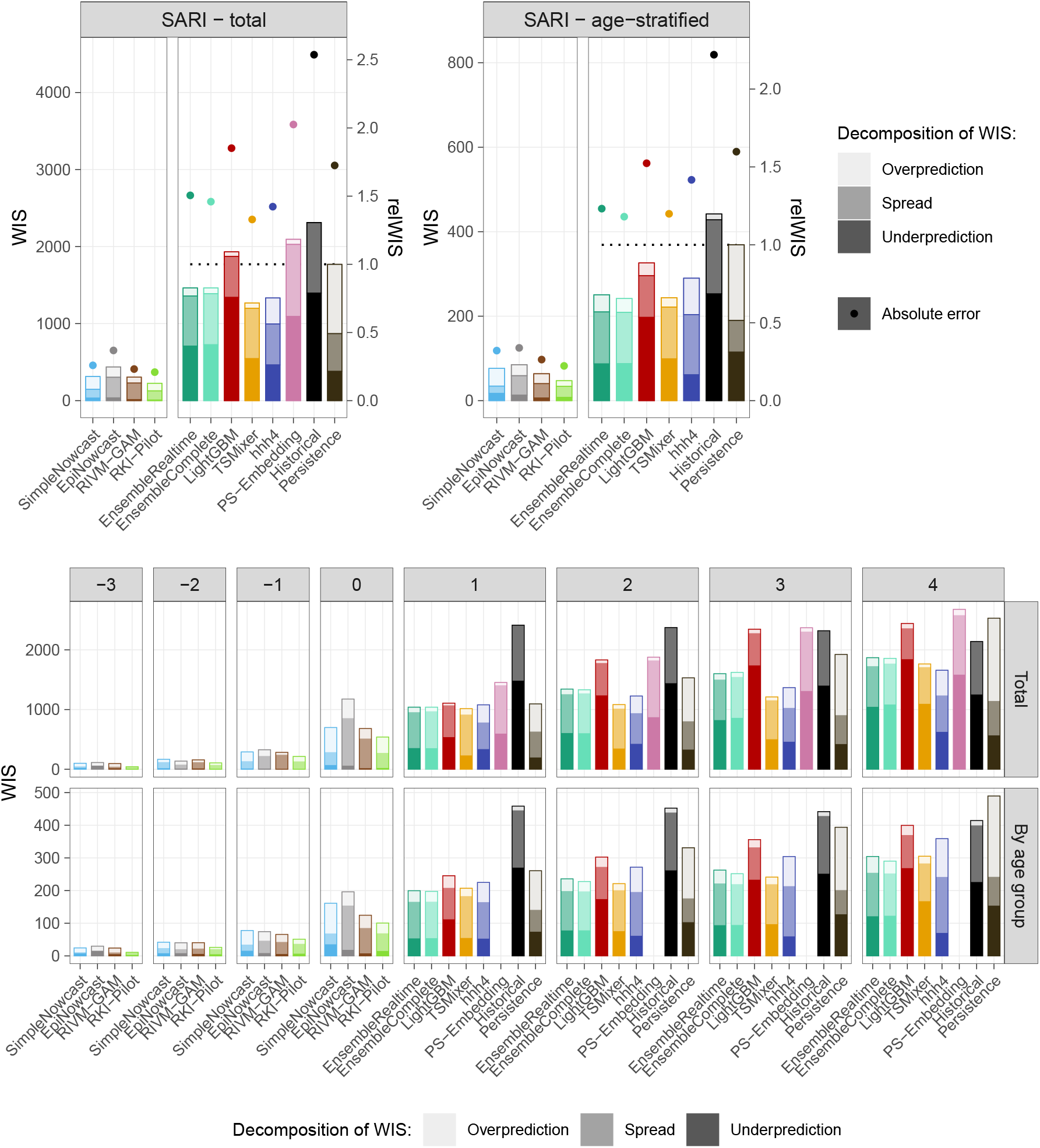
Summary of forecast performance for SARI in terms of weighted interval scores and absolute errors. See Figure 10 for details on plot elements. Average WIS values stratified by age group are available in Supplementary Figure S9. A numerical summary of the displayed values, along with statistical significance tests, is given in Supplementary Table S2.

The score decompositions are informative on the dispersion and biases of different models. For SARI, most models have pronounced underprediction components, while overprediction is almost absent. This is in line with the qualitative impression from Section 4.2.3, where underprediction during the high season was evident. A similar pattern, though less pronounced, also holds for the ARI indicator. The `LightGBM` model has noticeably smaller spread components than most other models, indicating more confident forecasting. For the ARI target, this makes it the best-performing model. For SARI, on the other hand, the model seems to suffer from overconfidence and performs much less favourably.

Results of permutation tests to check the statistical significance of performance differences relative to the `Persistence` baseline are reported in Supplementary Tables S1 and S2. Put shortly, for the total incidence counts, only the models `KIT-LightGBM` and `KIT-EnsembleComplete` for ARI achieve statistical significance at the 5% level, indicating that the 22 submission weeks are not sufficient to achieve good statistical power. For age-stratified predictions, where the number of issued forecasts is five (ARI) or six times (SARI) higher, most score differences are statistically significant.

When stratifying performance by horizon (bottom panels of both figures) it can be seen that at one week ahead, most models are roughly level with the `Persistence` baseline. This is less surprising for the ARI target, where the first half of the study period is roughly a plateau. For SARI, this reflects the fact that most models prematurely predicted a turnaround, leading them to obtain worse scores than the `Persistence` baseline over several weeks. For the same reasons, at horizon 4 weeks the models struggle to outperform the `Historical` baseline for ARI. Gains of the ensemble over both baselines simultaneously occur only at horizons 2 and 3 weeks ahead, and mainly for the SARI target.

Performance stratified by age group is shown in Supplementary Figures S8 and S9 for the two indicators. For these stratified indicators, the ensemble quite consistently outperforms the benchmark models, while among the member models `TSMixer` shows the most consistent performance.

Evaluations in terms of WIS applied to log-transformed incidence targets yield broadly similar results to the above. We show the results for total ARI and SARI incidences in Figure 12, while the corresponding results for age-stratified targets are available in Supplementary Figure S10.

**Figure 12:**
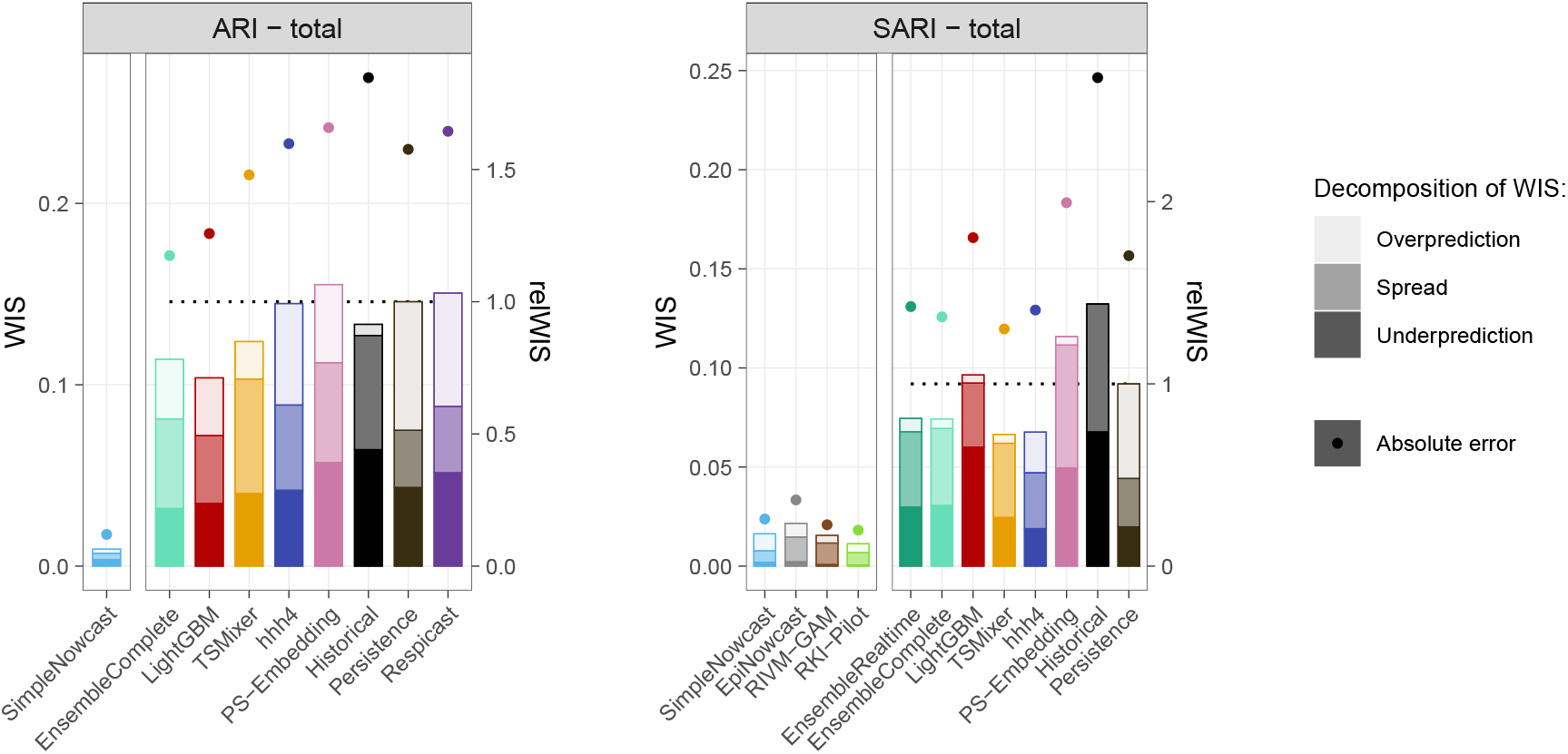
Summary of average WIS and absolute errors for log-transformed incidence targets. This figure shows results for total incidences, while the age-stratified results can be found in Supplementary Figure S10. Results of significance tests against the `Persistence` baseline are reported in Supplementary Tables S1 and S2.

## 5 Discussion

We provided an assessment of a prospective nowcasting and forecasting project for respiratory disease activity in Germany during the 2024/25 season, noting that part of the submissions needed to be completed retrospectively. Nowcasting generally showed satisfactory performance, although some models initially lacked mechanisms to appropriately handle the Christmas break. Averaged over submission weeks and horizons, the ensemble forecast and some member models achieved moderate improvements over two baseline models. Stratified by forecast horizon, however, most models showed performance close to a persistence model one week ahead, and close to a seasonal baseline 4 weeks ahead; forecast skill was thus mainly achieved at intermediate horizons.

The 2024/25 season represents the first operational season of the underlying *RESPINOW* Hub. As may be expected, this initial round exposed some weaknesses of models, that might be easily avoided in subsequent seasons – especially regarding the handling of the Christmas break effects in the ARI indicator. This aspect had not been examined closely enough as much of the preparatory analyses had focused on the SARI indicator (Wolffram et al., 2023; Günther et al., 2025). As seen for the `hhh4` model, the inclusion of indicator variables for the Christmas holidays can fix this problem.

Correct prediction of the peak timing turned out to be challenging, as has been remarked previously in the literature (Reich et al., 2019). As ARI and SARI are syndromic indicators, they represent a composite of epidemics from different pathogens, making it hard to anticipate the peak. Improvements may be feasible by considering stratified indicators, which for SARI have become available in the meantime. In our analysis we saw more success in predicting the peak timing for the pathogen-specific influenza, though not the RSV indicator. This observation, however, is based on a single model and season and should not be seen as a generalizable finding.

In our comparison we included the ensemble ARI forecasts provided by the ECDC effort *RespiCast* (Gozzi et al., 2026) for comparison. *RESPINOW* and *RespiCast* currently operate in parallel, but there is an ongoing exchange on best practices and potential synergies. The *RESPINOW* Hub covers a different set of indicators and is tailored to the data availability and empirical particularities of the German context. Notably, nowcasting, is not addressed in RespiCast, but an essential step in a forecasting pipeline for the German SARI indicator. Our national-level effort moreover lends itself better to including additional stratified time series by age group and more recently geographic region. A promising direction is the inclusion of pathogen-specific SARI counts, which have become available through Robert Koch Institute since the launch of *RESPINOW*.

A limitation of the presented study is that not all submissions were collected in real time, meaning there is some risk for hindsight bias. On an organizational level, we stress the challenges we encountered in ensuring regular participation. Despite the *RESPINOW* Hub being part of a modelling consortium for respiratory diseases, the number of models ultimately available remained moderate. This was partly because some available models turned out not to be suitable for well-calibrated short-term predictions. While one team that was external to the original consortium (RIVM) was among the most regular contributors, involving such additional teams proved difficult. Given the declining general focus on respiratory pathogens in infectious disease research and funding opportunities, these challenges were not entirely unexpected, and were among the motivations for embedding the project within the *RESPINOW* consortium.

Another limitation is the fact that all forecasting models depended on inputs from nowcasting steps, meaning that the observed performance at horizons 1 through 4 conflates performance of the nowcasting and forecasting models. We moreover note that most forecasting models (all `KIT` models) relied on the same nowcasting model (`KIT-simple_nowcast`). This is an intended feature of the Hub platform as such a modular approach considerably eases the development of forecasting models. It does, however, introduce a common source of error, which may hamper the diversity of the resulting ensemble.

Besides the scientific results, a major outcome of the *RESPINOW Hub* is its community building aspect, especially the strengthening of links between disease modelers from Germany’s federal public health agency Robert Koch Institute (RKI) and academic partner institutions. Notably, a nowcasting model developed and tested by RKI within the *RESPINOW* infrastructure subsequently became part of routine weekly reports (see Günther et al. 2025).

Some of the challenges in ensuring regular submissions may be alleviated by integrating new AI-based tools into Forecast Hub workflows. In a recent study, Martinson et al. (2026) demonstrate that coding agents are able to reliably implement forecasting models described in natural language, iteratively optimizing them for evaluation scores on a training window. Indeed, curated ensembles of several such models were able to outperform the US CDC *FluSight* ensemble (Mathis et al., 2024) under real-time conditions during the 2025/26 season. By leveraging such systems, it may be possible to maintain diverse sets of models with a small group of participating teams. However, besides questions of ethics and accountability, it is still unclear to which degree small academic teams with less expertise on agentic AI and more limited computing resources will be able to leverage these tools in practice.

## Data Availability

Replication data and codes for all shown analyses are available at https://github.com/KITmetricslab/RESPINOW-Hub-evaluation.

## Acknowledgements

This research was supported by the German Federal Ministry of Research, Technology and Space (BMFTR) via the project RESPINOW (grant number: HZI MV2021-012, KIT 031L0298J). JB acknowledges funding from the German Research Foundation (DFG), project 512483310. MS gratefully acknowledges funding via the Klaus Tschira Foundation and the Helmholtz grant COCAP (KA1-CO-10).

## Supplementary Material

### A Members of the RESPINOW Study Group

The following persons are members of the RESPINOW Study Group: Laura-Ines Boehler, Claudia Denkinger, Cornelia Gottschick, Manuela Harries, Torben Heinsohn, Olga Hovardovska, Veronika Jäger, André Karch, Carolina Klett-Tammen, Tyll Krüger, Patrick Marsall, Florian Marx, Rafael Mikolajczyk, Nicole Schneiderhan-Marra.

### B Additional technical details

#### B.1 Accounting for Christmas dip in the hhh4 model

Denoting weekly incidence counts by *X*_*t*_, the hhh4 model used here is defined as

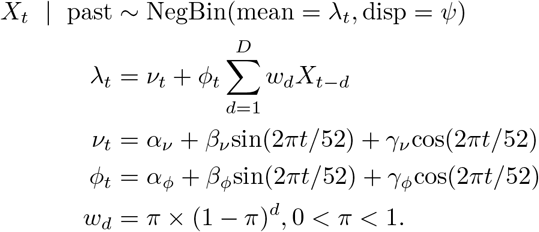

This model thus accounts for autocorrelation, (unimodal) yearly seasonality in magnitude and autocorrelation, as well as overdispersion. The weights *w*_*d*_ can be interpreted as the discrete-time generation time distribution. The model can be fitted using the R packages surveillance (Meyer et al., 2017) hhh4addon (Bracher and Held, 2022). The only addition needed to account for the Christmas break is to set

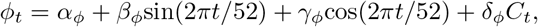

with *C*_*t*_ an indicator variable taking a value of one in calendar weeks 52 and 1 and zero otherwise.

#### B.2 Description of the PS-Embedding model

The `PS-Embedding_model` is based on a matching between historical incidence curves and the current seasonal course. Each incidence curve is translated to s series of tuples

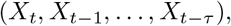

where in practice a value of *τ* = 3 is used. Figure S1 illustrates this idea in a two-dimensional phase diagrams (hence omitting two dimensions).

For now assuming that there are no reporting delays, the curve for the ongoing season ends in a given coordinate, highlighted by a red cross. Four nearest neighbour points are subsequently selected (yellow dots). For each of them, the further course of the respective season implies predictions at horizons one through four weeks ahead (shown by green dots for one week ahead). The overall point prediction results as the mean of these four trajectories.

The correction for reporting delays is based on a shift of the starting points (red crosses) estimated from historical data snapshots.

**Figure S1:**
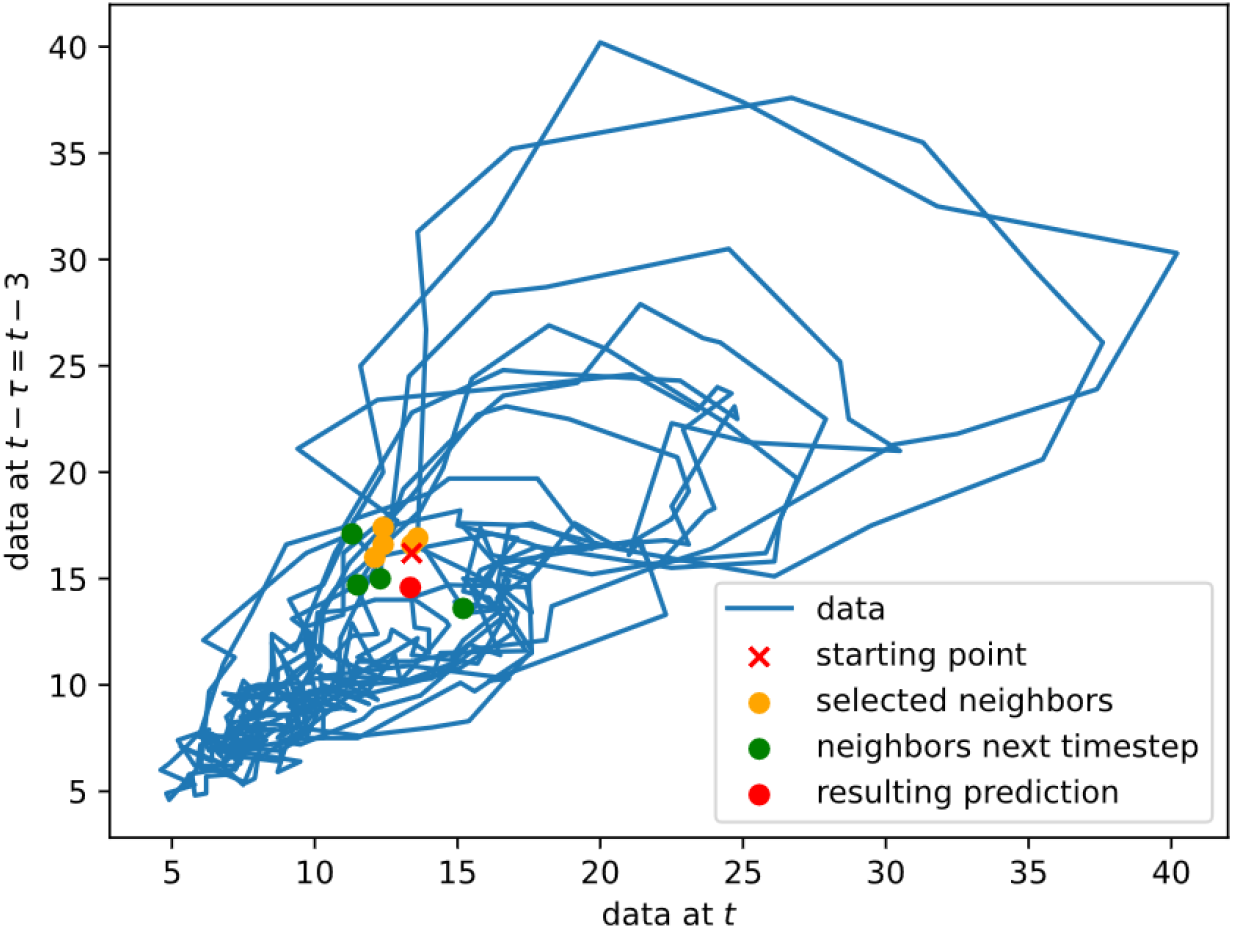
Graphical illustration of the `PS-Embedding` model.

### C Supplementary figures and tables

**Figure S2:**
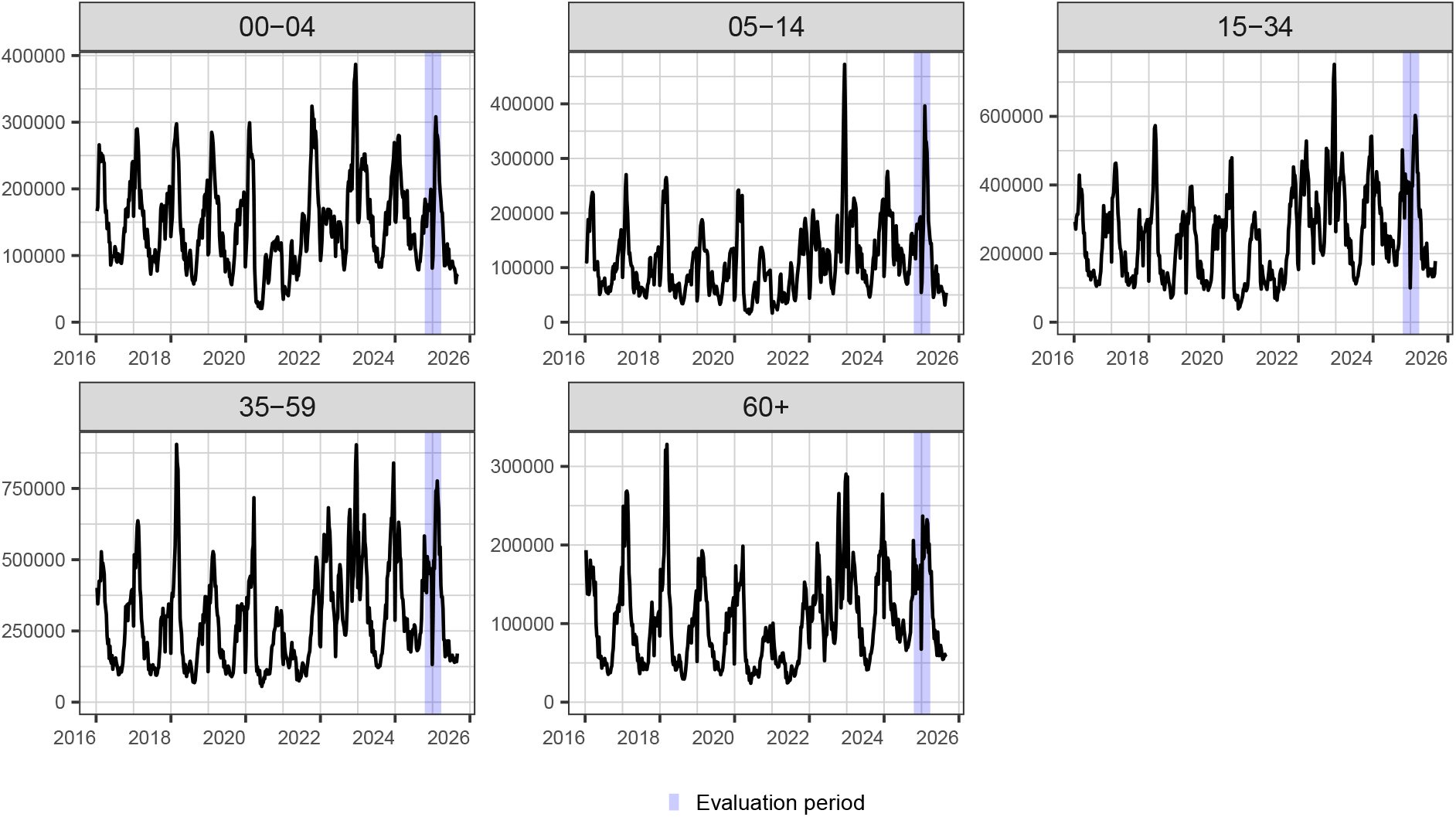
Age-stratified time series for the ARI indicator.

**Figure S3:**
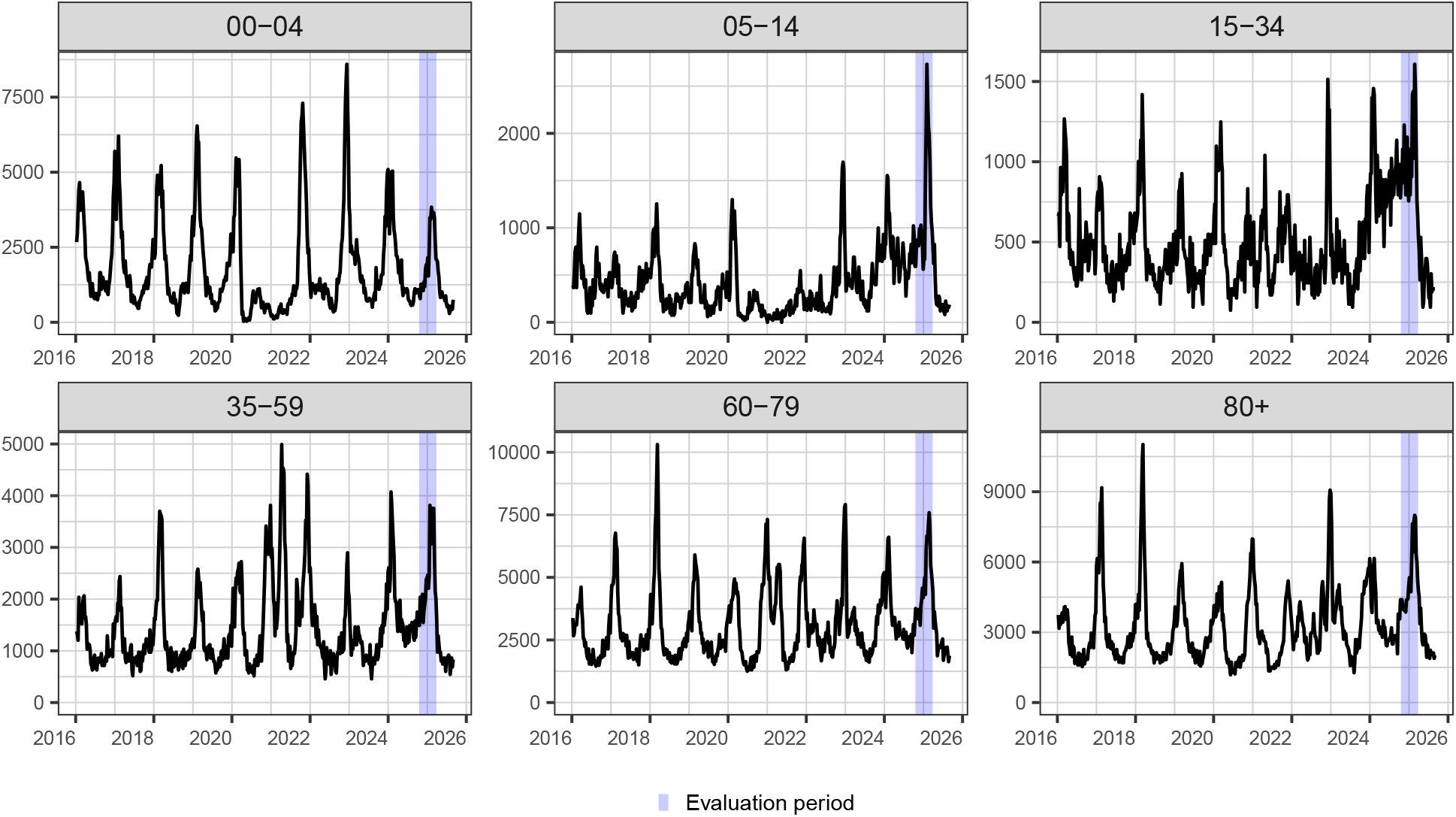
Age-stratified time series for the SARI indicator.

**Figure S4:**
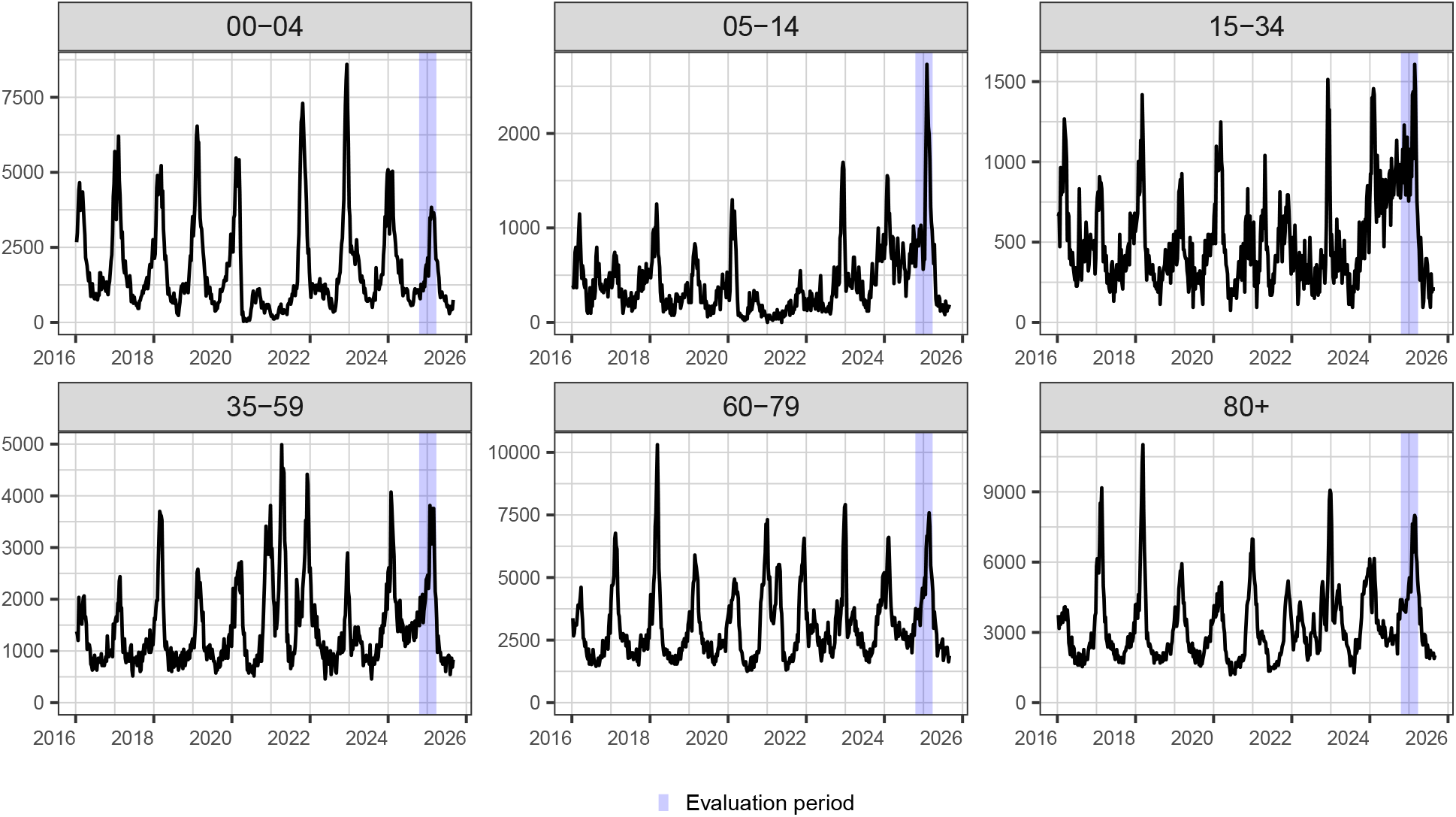
Age-stratified time series for the seasonal influenza indicator.

**Figure S5:**
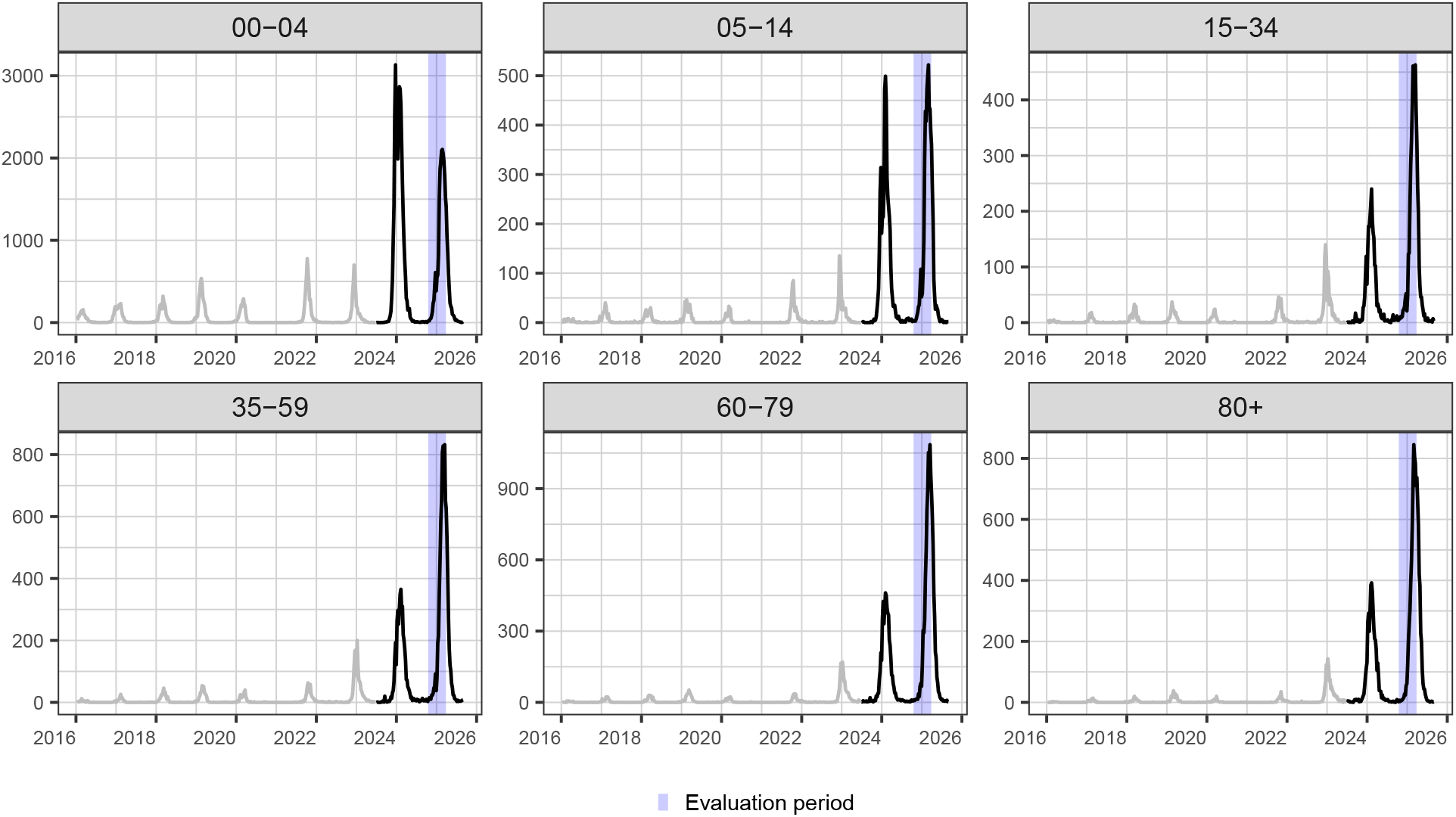
Age-stratified time series for the RSV indicator.

**Figure S6:**
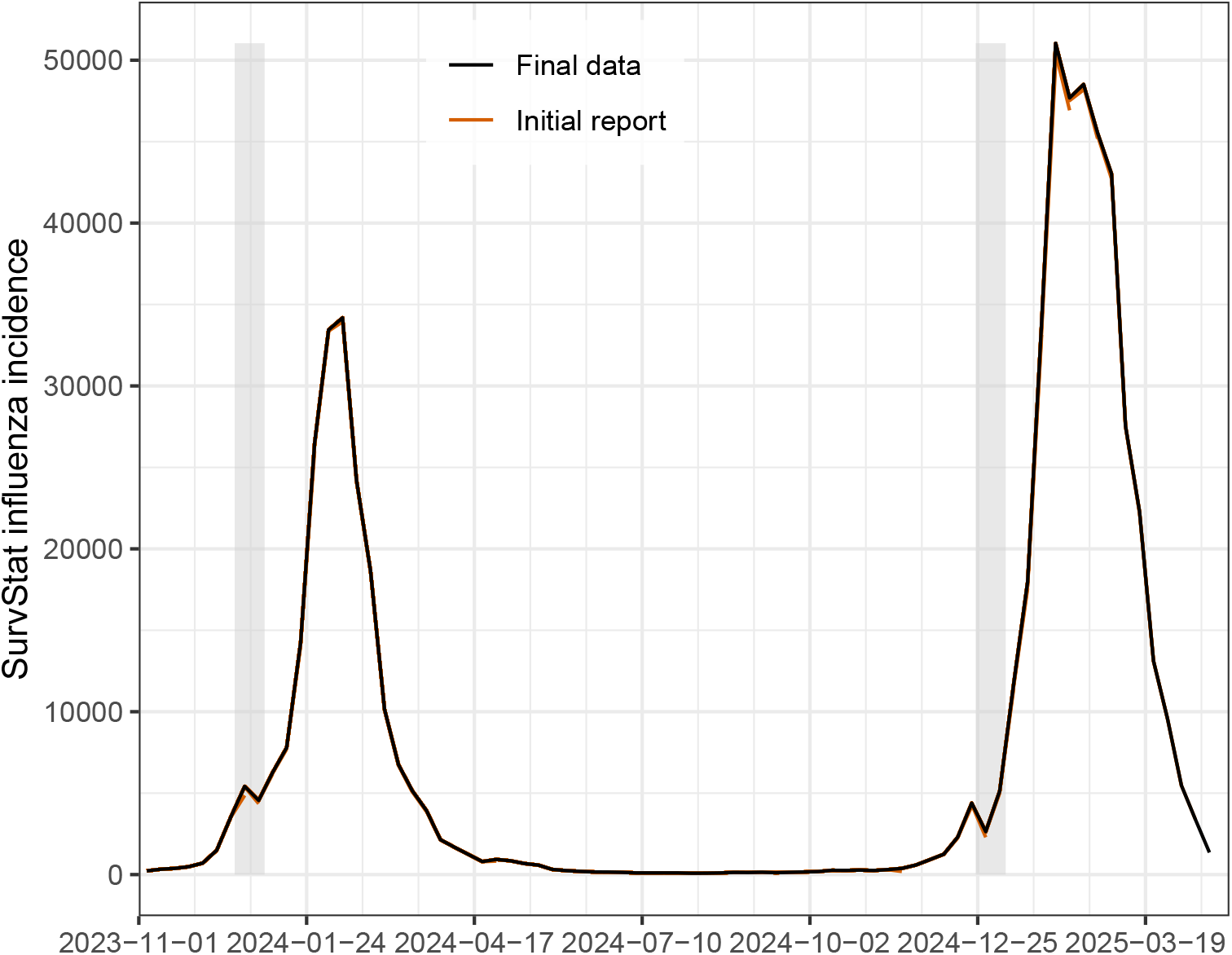
Revisions of SurvStat influenza data (paralleling Figure 2).

**Figure S7:**
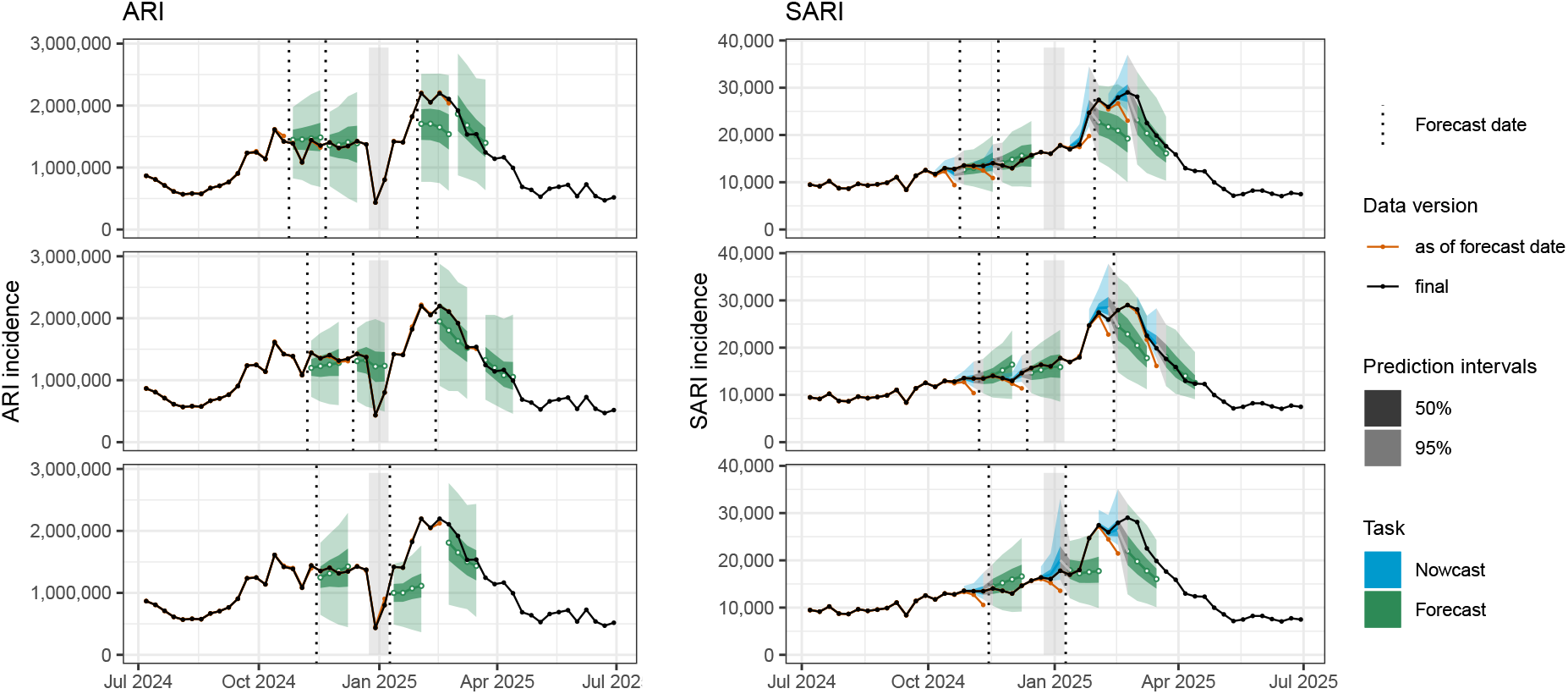
Selected nowcast and forecast trajectories for the ARI (left column) and SARI (right) indicators. This figure shows all weeks not contained in Figure 4 from the main manuscript.

**Figure S8:**
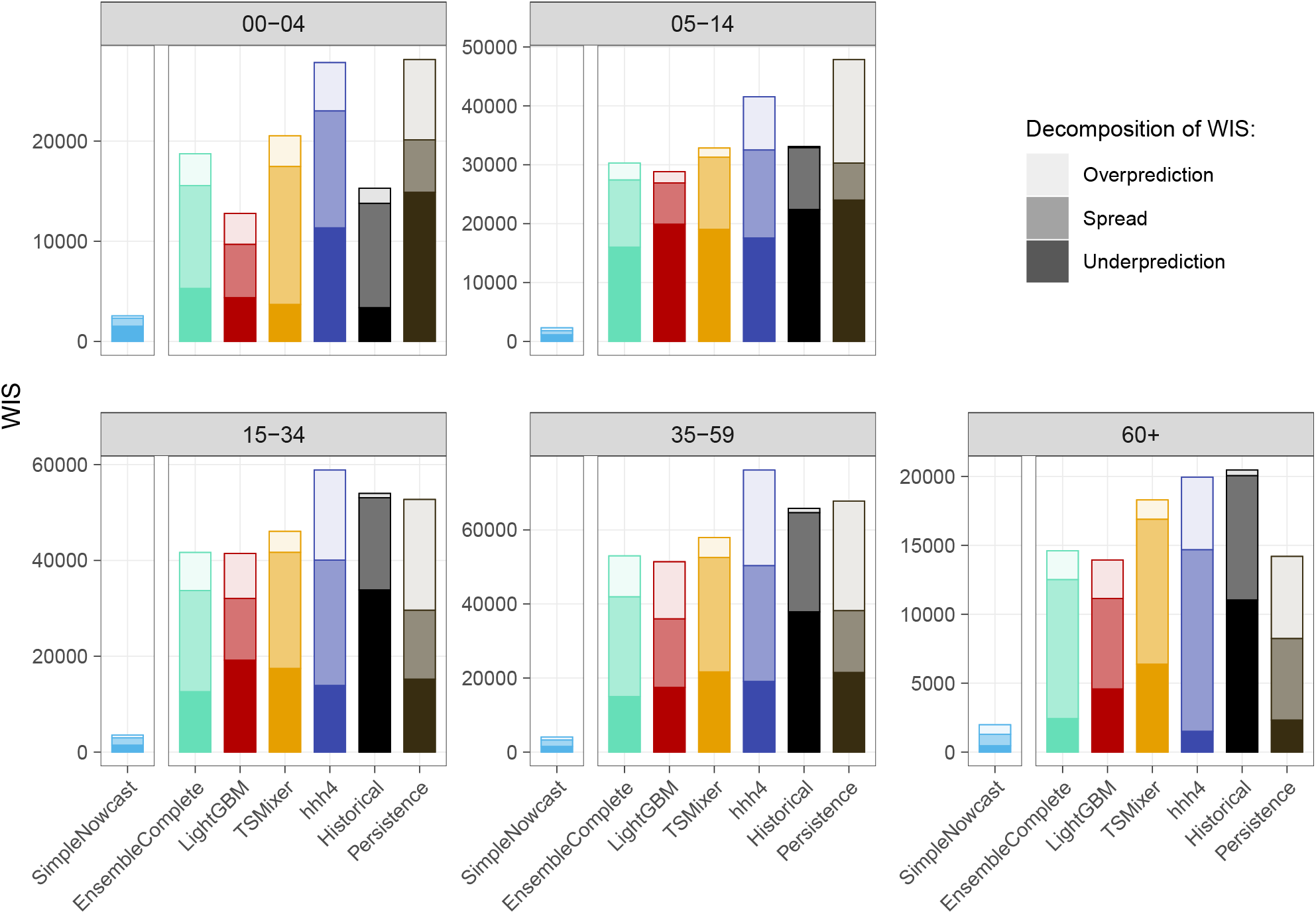
Average WIS values for age group-wise ARI incidences, by age group.

**Figure S9:**
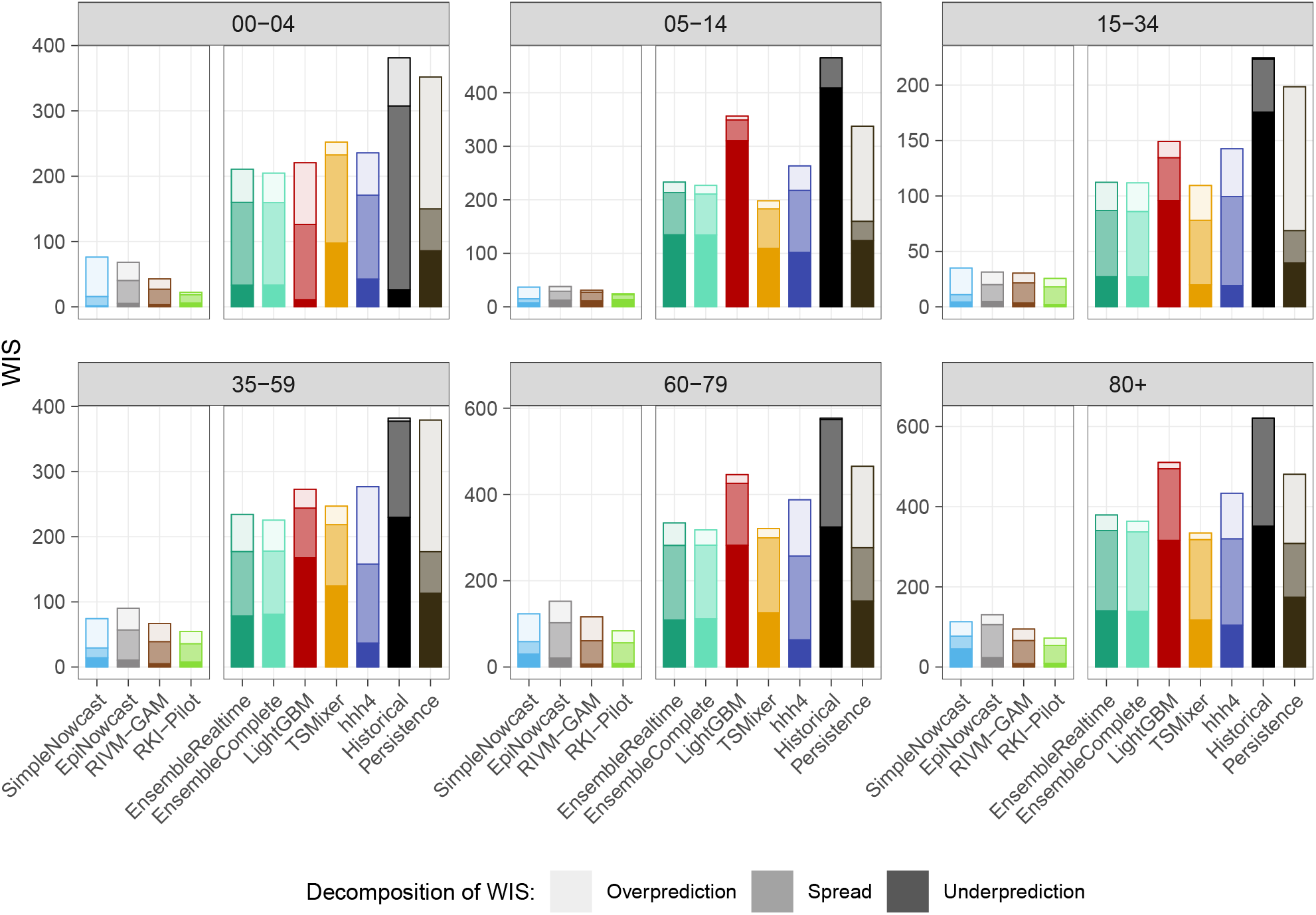
Average WIS values for age group-wise SARI incidences, by age group.

**Table S1:**
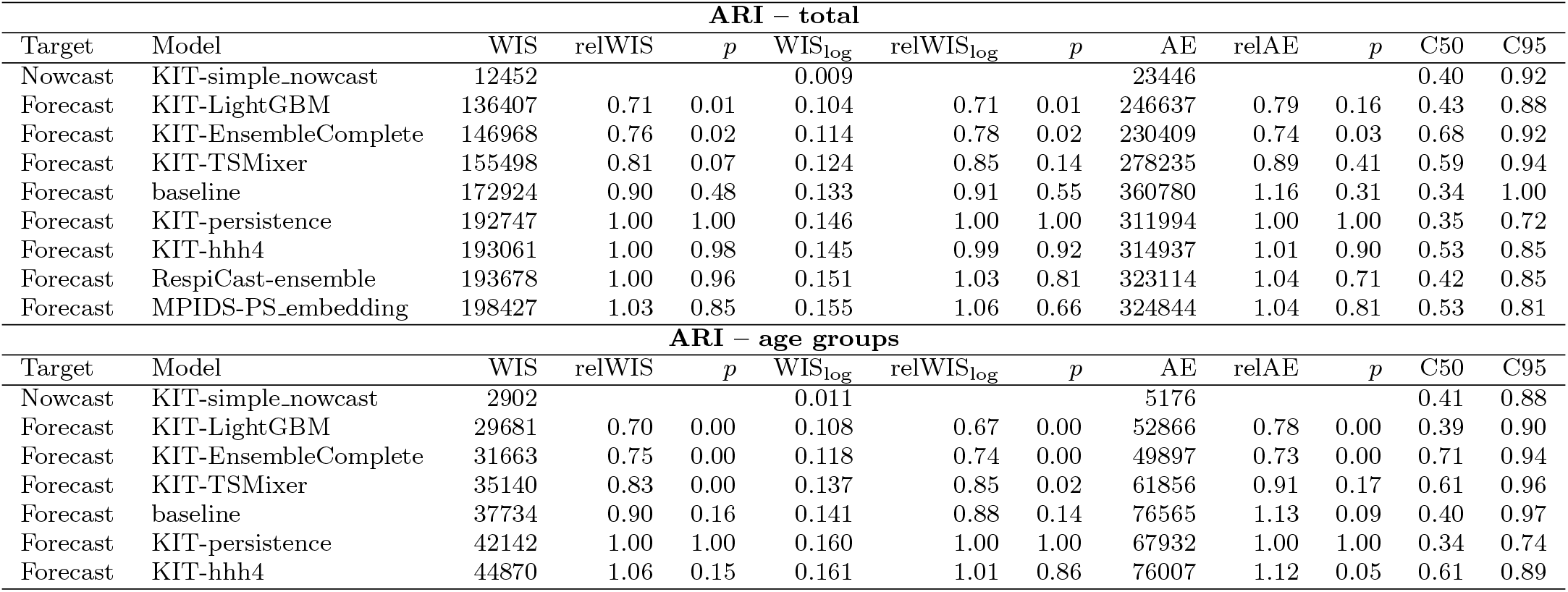
Summary table on model performance for the ARI data. All relative scores are given with respect to the `KIT-persistence` baseline. WIS_log_ is the weighted interval score applied to the transformed target log(incidence count + 1). C50 and C95 are coverage fractions of the 95% and 50% prediction intervals. *p*-values refer to permutation tests comparing each model to the `Persistence` baseline.

| **ARI – total** |  |  |  |  |  |  |  |  |  |  |  |  |
|---|---|---|---|---|---|---|---|---|---|---|---|---|
| Target | Model | WIS | relWIS | *p* | WIS<sub>log</sub> | relWIS<sub>log</sub> | *p* | AE | relAE | *p* | C50 | C95 |
| Nowcast | KIT-simple_nowcast | 12452 |  |  | 0.009 |  |  | 23446 |  |  | 0.40 | 0.92 |
| Forecast | KIT-LightGBM | 136407 | 0.71 | 0.01 | 0.104 | 0.71 | 0.01 | 246637 | 0.79 | 0.16 | 0.43 | 0.88 |
| Forecast | KIT-EnsembleComplete | 146968 | 0.76 | 0.02 | 0.114 | 0.78 | 0.02 | 230409 | 0.74 | 0.03 | 0.68 | 0.92 |
| Forecast | KIT-TSMixer | 155498 | 0.81 | 0.07 | 0.124 | 0.85 | 0.14 | 278235 | 0.89 | 0.41 | 0.59 | 0.94 |
| Forecast | baseline | 172924 | 0.90 | 0.48 | 0.133 | 0.91 | 0.55 | 360780 | 1.16 | 0.31 | 0.34 | 1.00 |
| Forecast | KIT-persistence | 192747 | 1.00 | 1.00 | 0.146 | 1.00 | 1.00 | 311994 | 1.00 | 1.00 | 0.35 | 0.72 |
| Forecast | KIT-hhh4 | 193061 | 1.00 | 0.98 | 0.145 | 0.99 | 0.92 | 314937 | 1.01 | 0.90 | 0.53 | 0.85 |
| Forecast | RespiCast-ensemble | 193678 | 1.00 | 0.96 | 0.151 | 1.03 | 0.81 | 323114 | 1.04 | 0.71 | 0.42 | 0.85 |
| Forecast | MPIDS-PS_embedding | 198427 | 1.03 | 0.85 | 0.155 | 1.06 | 0.66 | 324844 | 1.04 | 0.81 | 0.53 | 0.81 |
| **ARI – age groups** |  |  |  |  |  |  |  |  |  |  |  |  |
| Target | Model | WIS | relWIS | *p* | WIS<sub>log</sub> | relWIS<sub>log</sub> | *p* | AE | relAE | *p* | C50 | C95 |
| Nowcast | KIT-simple_nowcast | 2902 |  |  | 0.011 |  |  | 5176 |  |  | 0.41 | 0.88 |
| Forecast | KIT-LightGBM | 29681 | 0.70 | 0.00 | 0.108 | 0.67 | 0.00 | 52866 | 0.78 | 0.00 | 0.39 | 0.90 |
| Forecast | KIT-EnsembleComplete | 31663 | 0.75 | 0.00 | 0.118 | 0.74 | 0.00 | 49897 | 0.73 | 0.00 | 0.71 | 0.94 |
| Forecast | KIT-TSMixer | 35140 | 0.83 | 0.00 | 0.137 | 0.85 | 0.02 | 61856 | 0.91 | 0.17 | 0.61 | 0.96 |
| Forecast | baseline | 37734 | 0.90 | 0.16 | 0.141 | 0.88 | 0.14 | 76565 | 1.13 | 0.09 | 0.40 | 0.97 |
| Forecast | KIT-persistence | 42142 | 1.00 | 1.00 | 0.160 | 1.00 | 1.00 | 67932 | 1.00 | 1.00 | 0.34 | 0.74 |
| Forecast | KIT-hhh4 | 44870 | 1.06 | 0.15 | 0.161 | 1.01 | 0.86 | 76007 | 1.12 | 0.05 | 0.61 | 0.89 |

**Figure S10:**
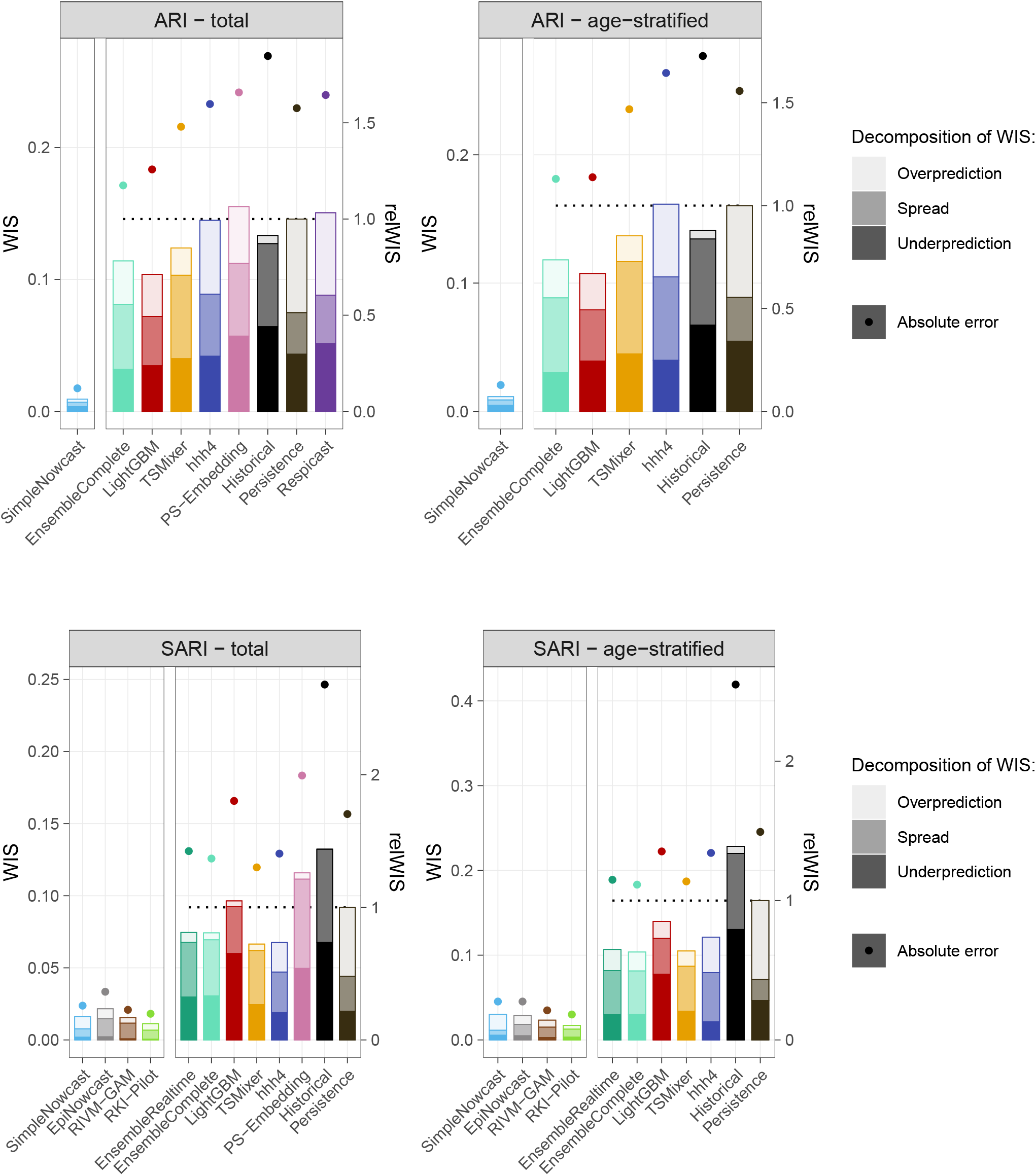
Average WIS values for log-transformed incidences for total and age-stratified targets.

**Figure S11:**
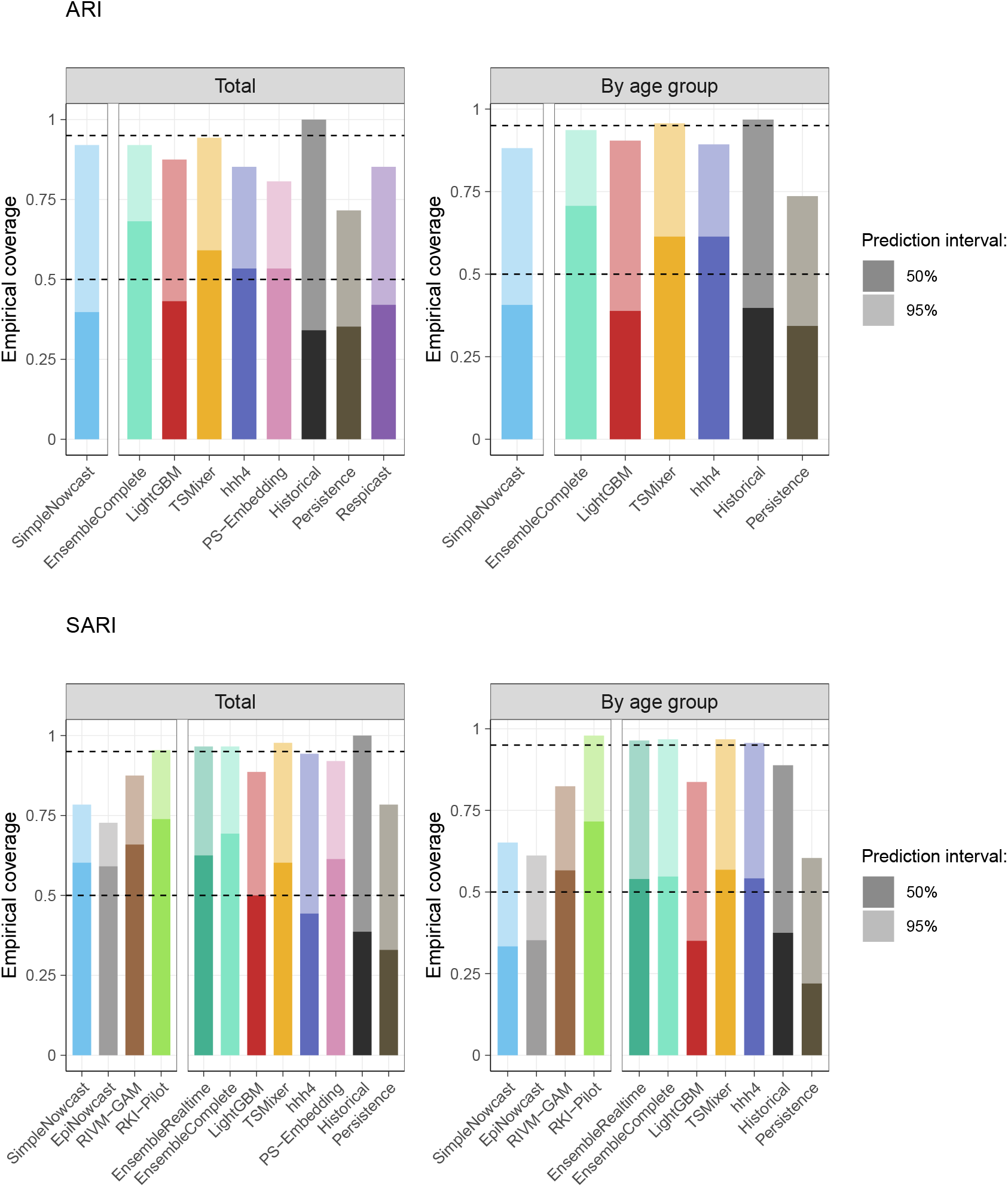
Coverage of 50% and 95% prediction intervals for the ARI (top) and SARI (bottom) targets. Fractions are averaged across forecast horizons and weeks. The left column shows results for the total incidence time series, the right for age-stratified nowcasts and forecasts. Within each panel, results for nowcasting and forecasting models are shown separately.

**Figure S12:**
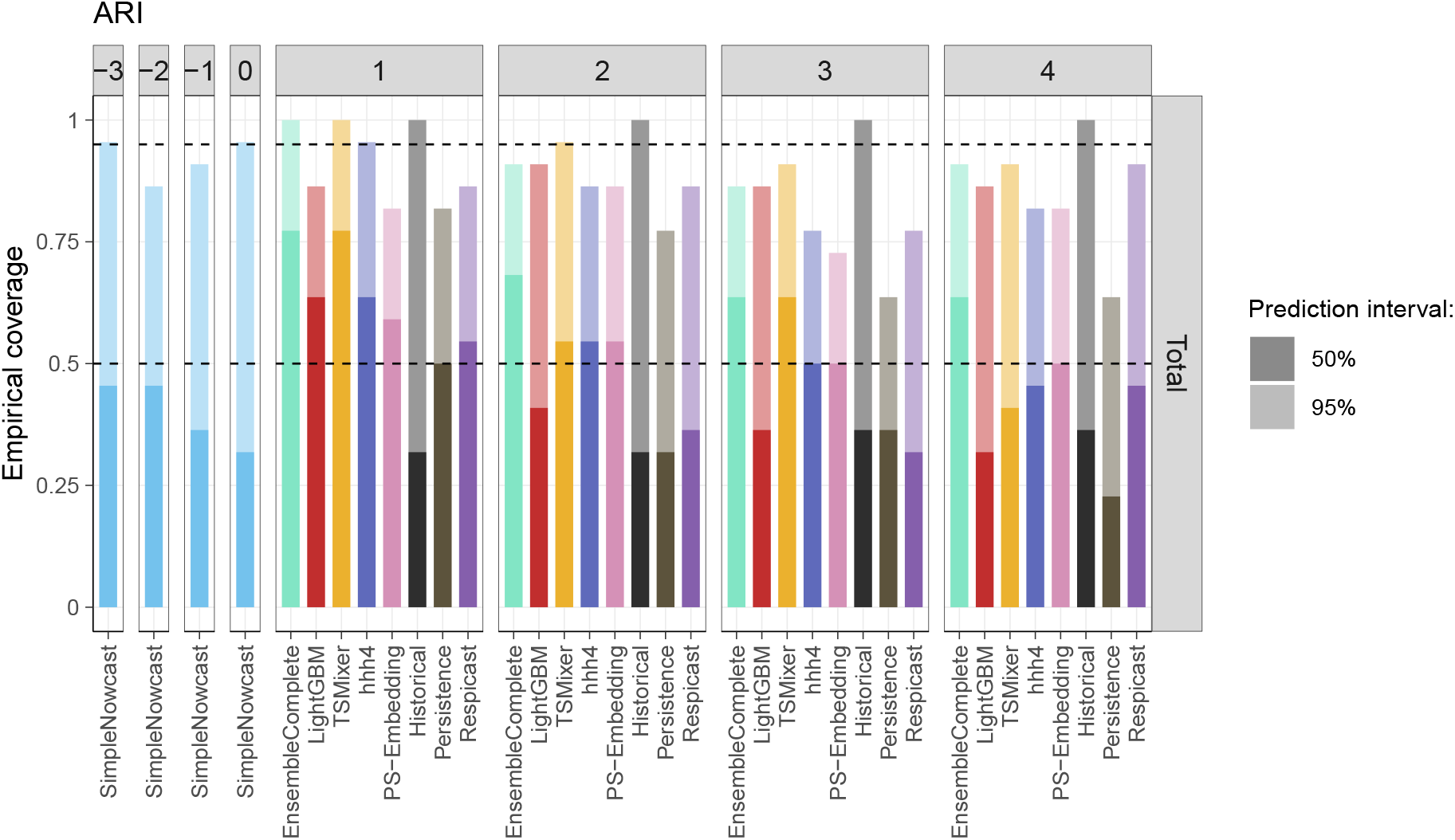
Prediction interval coverage by forecast horizon for the ARI indicator (complementing Figure 9).

**Figure S13:**
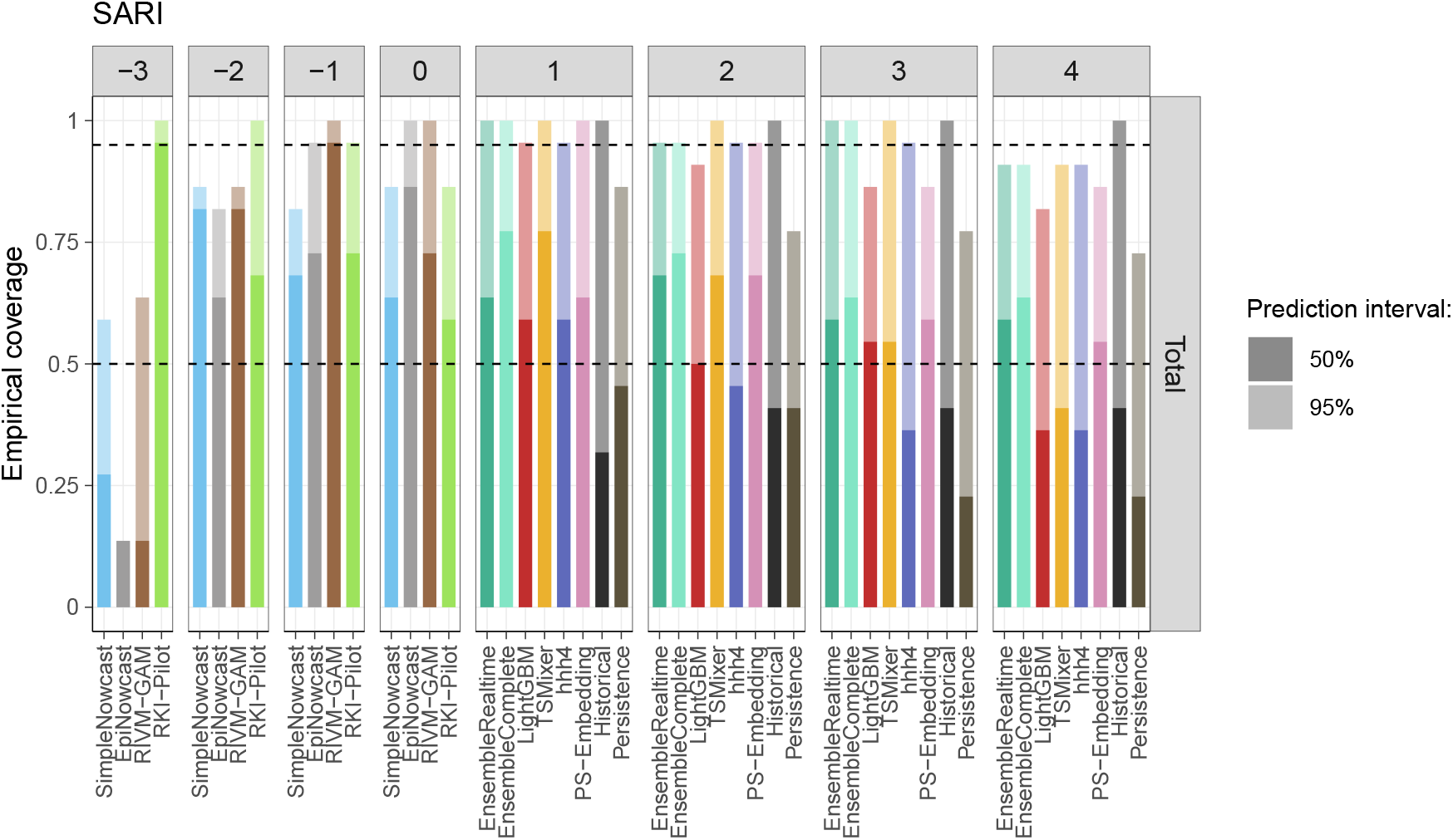
Prediction interval coverage by forecast horizon for the SARI indicator (complementing Figure 9).

**Table S2:**
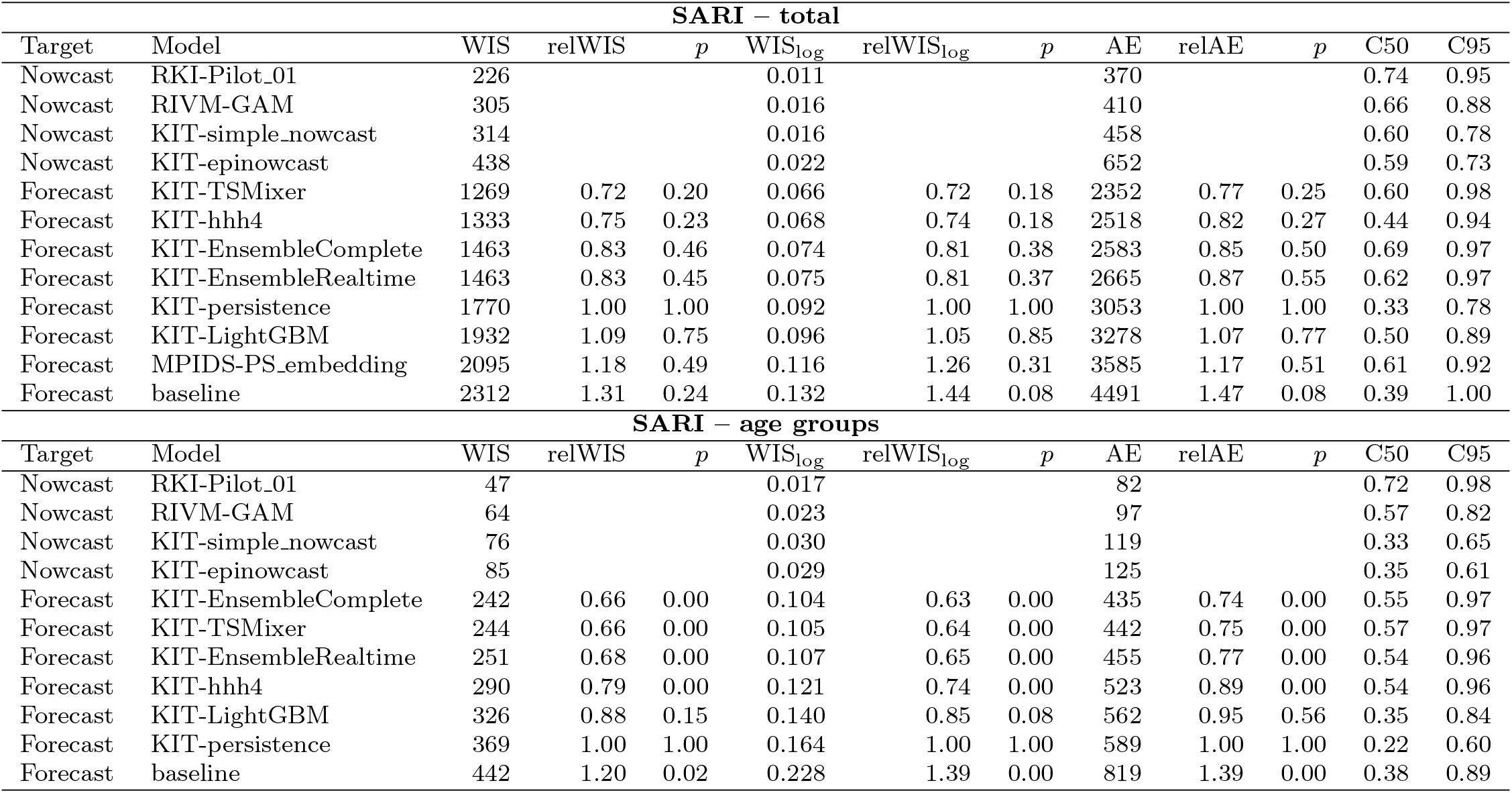
Summary table on model performance for the SARI data. All relative scores are given with respect to the `KIT-persistence` baseline. WIS_log_ is the weighted interval score applied to the transformed target log(incidence count + 1). C50 and C95 are coverage fractions of the 95% and 50% prediction intervals. *p*-values refer to permutation tests comparing each model to the `Persistence` baseline

| **SARI – total** |  |  |  |  |  |  |  |  |  |  |  |  |
|---|---|---|---|---|---|---|---|---|---|---|---|---|
| Target | Model | WIS | relWIS | *p* | WIS<sub>log</sub> | relWIS<sub>log</sub> | *p* | AE | relAE | *p* | C50 | C95 |
| Nowcast | RKI-Pilot_01 | 226 |  |  | 0.011 |  |  | 370 |  |  | 0.74 | 0.95 |
| Nowcast | RIVM-GAM | 305 |  |  | 0.016 |  |  | 410 |  |  | 0.66 | 0.88 |
| Nowcast | KIT-simple_nowcast | 314 |  |  | 0.016 |  |  | 458 |  |  | 0.60 | 0.78 |
| Nowcast | KIT-epinowcast | 438 |  |  | 0.022 |  |  | 652 |  |  | 0.59 | 0.73 |
| Forecast | KIT-TSMixer | 1269 | 0.72 | 0.20 | 0.066 | 0.72 | 0.18 | 2352 | 0.77 | 0.25 | 0.60 | 0.98 |
| Forecast | KIT-hhh4 | 1333 | 0.75 | 0.23 | 0.068 | 0.74 | 0.18 | 2518 | 0.82 | 0.27 | 0.44 | 0.94 |
| Forecast | KIT-EnsembleComplete | 1463 | 0.83 | 0.46 | 0.074 | 0.81 | 0.38 | 2583 | 0.85 | 0.50 | 0.69 | 0.97 |
| Forecast | KIT-EnsembleRealtime | 1463 | 0.83 | 0.45 | 0.075 | 0.81 | 0.37 | 2665 | 0.87 | 0.55 | 0.62 | 0.97 |
| Forecast | KIT-persistence | 1770 | 1.00 | 1.00 | 0.092 | 1.00 | 1.00 | 3053 | 1.00 | 1.00 | 0.33 | 0.78 |
| Forecast | KIT-LightGBM | 1932 | 1.09 | 0.75 | 0.096 | 1.05 | 0.85 | 3278 | 1.07 | 0.77 | 0.50 | 0.89 |
| Forecast | MPIDS-PS_embedding | 2095 | 1.18 | 0.49 | 0.116 | 1.26 | 0.31 | 3585 | 1.17 | 0.51 | 0.61 | 0.92 |
| Forecast | baseline | 2312 | 1.31 | 0.24 | 0.132 | 1.44 | 0.08 | 4491 | 1.47 | 0.08 | 0.39 | 1.00 |
| **SARI – age groups** |  |  |  |  |  |  |  |  |  |  |  |  |
| Target | Model | WIS | relWIS | *p* | WIS<sub>log</sub> | relWIS<sub>log</sub> | *p* | AE | relAE | *p* | C50 | C95 |
| Nowcast | RKI-Pilot_01 | 47 |  |  | 0.017 |  |  | 82 |  |  | 0.72 | 0.98 |
| Nowcast | RIVM-GAM | 64 |  |  | 0.023 |  |  | 97 |  |  | 0.57 | 0.82 |
| Nowcast | KIT-simple_nowcast | 76 |  |  | 0.030 |  |  | 119 |  |  | 0.33 | 0.65 |
| Nowcast | KIT-epinowcast | 85 |  |  | 0.029 |  |  | 125 |  |  | 0.35 | 0.61 |
| Forecast | KIT-EnsembleComplete | 242 | 0.66 | 0.00 | 0.104 | 0.63 | 0.00 | 435 | 0.74 | 0.00 | 0.55 | 0.97 |
| Forecast | KIT-TSMixer | 244 | 0.66 | 0.00 | 0.105 | 0.64 | 0.00 | 442 | 0.75 | 0.00 | 0.57 | 0.97 |
| Forecast | KIT-EnsembleRealtime | 251 | 0.68 | 0.00 | 0.107 | 0.65 | 0.00 | 455 | 0.77 | 0.00 | 0.54 | 0.96 |
| Forecast | KIT-hhh4 | 290 | 0.79 | 0.00 | 0.121 | 0.74 | 0.00 | 523 | 0.89 | 0.00 | 0.54 | 0.96 |
| Forecast | KIT-LightGBM | 326 | 0.88 | 0.15 | 0.140 | 0.85 | 0.08 | 562 | 0.95 | 0.56 | 0.35 | 0.84 |
| Forecast | KIT-persistence | 369 | 1.00 | 1.00 | 0.164 | 1.00 | 1.00 | 589 | 1.00 | 1.00 | 0.22 | 0.60 |
| Forecast | baseline | 442 | 1.20 | 0.02 | 0.228 | 1.39 | 0.00 | 819 | 1.39 | 0.00 | 0.38 | 0.89 |

